# Characteristics, outcomes, and mortality amongst 133,589 patients with prevalent autoimmune diseases diagnosed with, and 48,418 hospitalised for COVID-19: a multinational distributed network cohort analysis

**DOI:** 10.1101/2020.11.24.20236802

**Authors:** Eng Hooi Tan, Anthony G. Sena, Albert Prats-Uribe, Seng Chan You, Waheed-Ul-Rahman Ahmed, Kristin Kostka, Christian Reich, Scott L. Duvall, Kristine E. Lynch, Michael E. Matheny, Talita Duarte-Salles, Sergio Fernandez Bertolin, George Hripcsak, Karthik Natarajan, Thomas Falconer, Matthew Spotnitz, Anna Ostropolets, Clair Blacketer, Thamir M Alshammari, Heba Alghoul, Osaid Alser, Jennifer C.E. Lane, Dalia M Dawoud, Karishma Shah, Yue Yang, Lin Zhang, Carlos Areia, Asieh Golozar, Martina Relcade, Paula Casajust, Jitendra Jonnagaddala, Vignesh Subbian, David Vizcaya, Lana YH Lai, Fredrik Nyberg, Daniel R Morales, Jose D. Posada, Nigam H. Shah, Mengchun Gong, Arani Vivekanantham, Aaron Abend, Evan P Minty, Marc Suchard, Peter Rijnbeek, Patrick B Ryan, Daniel Prieto-Alhambra

## Abstract

**Objective:** Patients with autoimmune diseases were advised to shield to avoid COVID-19, but information on their prognosis is lacking. We characterised 30-day outcomes and mortality after hospitalisation with COVID-19 among patients with prevalent autoimmune diseases, and compared outcomes after hospital admissions among similar patients with seasonal influenza.

**Design:** Multinational network cohort study

**Setting:** Electronic health records data from Columbia University Irving Medical Center (CUIMC) (NYC, United States [US]), Optum [US], Department of Veterans Affairs (VA) (US), Information System for Research in Primary Care-Hospitalisation Linked Data (SIDIAP-H) (Spain), and claims data from IQVIA Open Claims (US) and Health Insurance and Review Assessment (HIRA) (South Korea).

**Participants:** All patients with prevalent autoimmune diseases, diagnosed and/or hospitalised between January and June 2020 with COVID-19, and similar patients hospitalised with influenza in 2017-2018 were included.

**Main outcome measures:** 30-day complications during hospitalisation and death

**Results:** We studied 133,589 patients diagnosed and 48,418 hospitalised with COVID-19 with prevalent autoimmune diseases. The majority of participants were female (60.5% to 65.9%) and aged ≥50 years. The most prevalent autoimmune conditions were psoriasis (3.5 to 32.5%), rheumatoid arthritis (3.9 to 18.9%), and vasculitis (3.3 to 17.6%). Amongst hospitalised patients, Type 1 diabetes was the most common autoimmune condition (4.8% to 7.5%) in US databases, rheumatoid arthritis in HIRA (18.9%), and psoriasis in SIDIAP-H (26.4%).

Compared to 70,660 hospitalised with influenza, those admitted with COVID-19 had more respiratory complications including pneumonia and acute respiratory distress syndrome, and higher 30-day mortality (2.2% to 4.3% versus 6.3% to 24.6%).

**Conclusions:** Patients with autoimmune diseases had high rates of respiratory complications and 30-day mortality following a hospitalization with COVID-19. Compared to influenza, COVID-19 is a more severe disease, leading to more complications and higher mortality. Future studies should investigate predictors of poor outcomes in COVID-19 patients with autoimmune diseases.

**What is already known about this topic:**

- Patients with autoimmune conditions may be at increased risk of COVID-19 infection andcomplications.
- There is a paucity of evidence characterising the outcomes of hospitalised COVID-19 patients with prevalent autoimmune conditions.

**What this study adds:**

- Most people with autoimmune diseases who required hospitalisation for COVID-19 were women, aged 50 years or older, and had substantial previous comorbidities.
- Patients who were hospitalised with COVID-19 and had prevalent autoimmune diseases had higher prevalence of hypertension, chronic kidney disease, heart disease, and Type 2 diabetes as compared to those with prevalent autoimmune diseases who were diagnosed with COVID-19.
- A variable proportion of 6% to 25% across data sources died within one month of hospitalisation with COVID-19 and prevalent autoimmune diseases.
- For people with autoimmune diseases, COVID-19 hospitalisation was associated with worse outcomes and 30-day mortality compared to admission with influenza in the 2017-2018 season.

## Introduction

Millions of people have been diagnosed, and hundreds of thousands have died from coronavirus disease 2019 (COVID-19) globally. (1) There is concern that patients with autoimmune diseases are at an increased risk of infection and complications, exacerbated by the nature of their disease and/or the use of immunosuppressive therapies.(2) In addition, systemic inflammation is present in many autoimmune diseases (3), leading to an increased risk of cardiovascular (3-5) and thromboembolic disease (6-8), have also been recently reported to be associated with COVID-19. In patients infected with COVID-19, worse outcomes such as hospitalisation, requiring intensive services, and death may be associated with a pro-inflammatory cytokine storm.(9-11) Currently identified general risk factors for COVID-19 hospitalisation include systemic autoimmune diseases amongst other comorbidities.(12, 13)

As having autoimmune diseases is a recognised risk factor for COVID-19 related complications (2), public health authorities around the world have advised mitigation strategies for those at risk. In the absence of a vaccine and a scarcity of proven therapeutic options, non-pharmacological measures such as shielding, case isolation, strict hand hygiene, and social distancing are key measures to protect this vulnerable group of patients.(14, 15) Thus far, characterisation studies about COVID-19 infection in people with autoimmune conditions have been limited in sample size and mostly region-specific.(12, 13, 16-19) As such, COVID-19 outcomes among people with autoimmune conditions remain poorly understood.

With the ongoing threat of COVID-19, clinical understanding of the characteristics and prognosis of patients with autoimmune conditions will facilitate the management of care for this group of patients. Given the paucity of evidence, our study aimed to describe the patients’ socio-demographics, comorbidities, and 30-day complications and mortality amongst patients with prevalent autoimmune conditions hospitalised and COVID-19 across North America, Europe, and Asia. In addition, we compared their health outcomes and mortality with those seen in patients with autoimmune diseases hospitalised with seasonal influenza in the previous years.

## Methods

### Study design and data sources

We conducted a multinational network retrospective cohort study as part of the Characterizing Health Associated Risks, and Your Baseline Disease In SARS-COV-2 (CHARYBDIS) protocol.(20) At time of publication, there were 18 databases contributing to CHARYBDIS. All data were standardized to the Observational Medical Outcomes Partnership (OMOP) Common Data Model (CDM)(21), which allowed a federated network analysis without sharing patient-level data. In this study, we selected databases with more than 140 patients meeting our inclusion criteria to secure sufficient precision with a confidence interval width of +/-5% in the study of the prevalence of a previous condition or 30-day risk of an outcome affecting 10% of the study population.

We included six data sources from the US, Spain, and South Korea, including hospital out- and inpatient electronic health records (EHR) from Columbia University Irving Medical Center (CUIMC) US, Optum (Optum EHR) (US), Department of Veterans Affairs (VA-OMOP) (US), primary care EHR linked to hospital admissions data from the Information System for Research in Primary Care-Hospitalisation Linked Data (SIDIAP-H) (Spain)(22), and health claims from IQVIA Open Claims (US) and Health Insurance and Review Assessment (HIRA) (South Korea).(23) A flowchart of the databases included and excluded of those available in the network is shown in Supplementary Figure 1, and a detailed description of the included databases can be found in Appendix 1.

**Figure 1.**
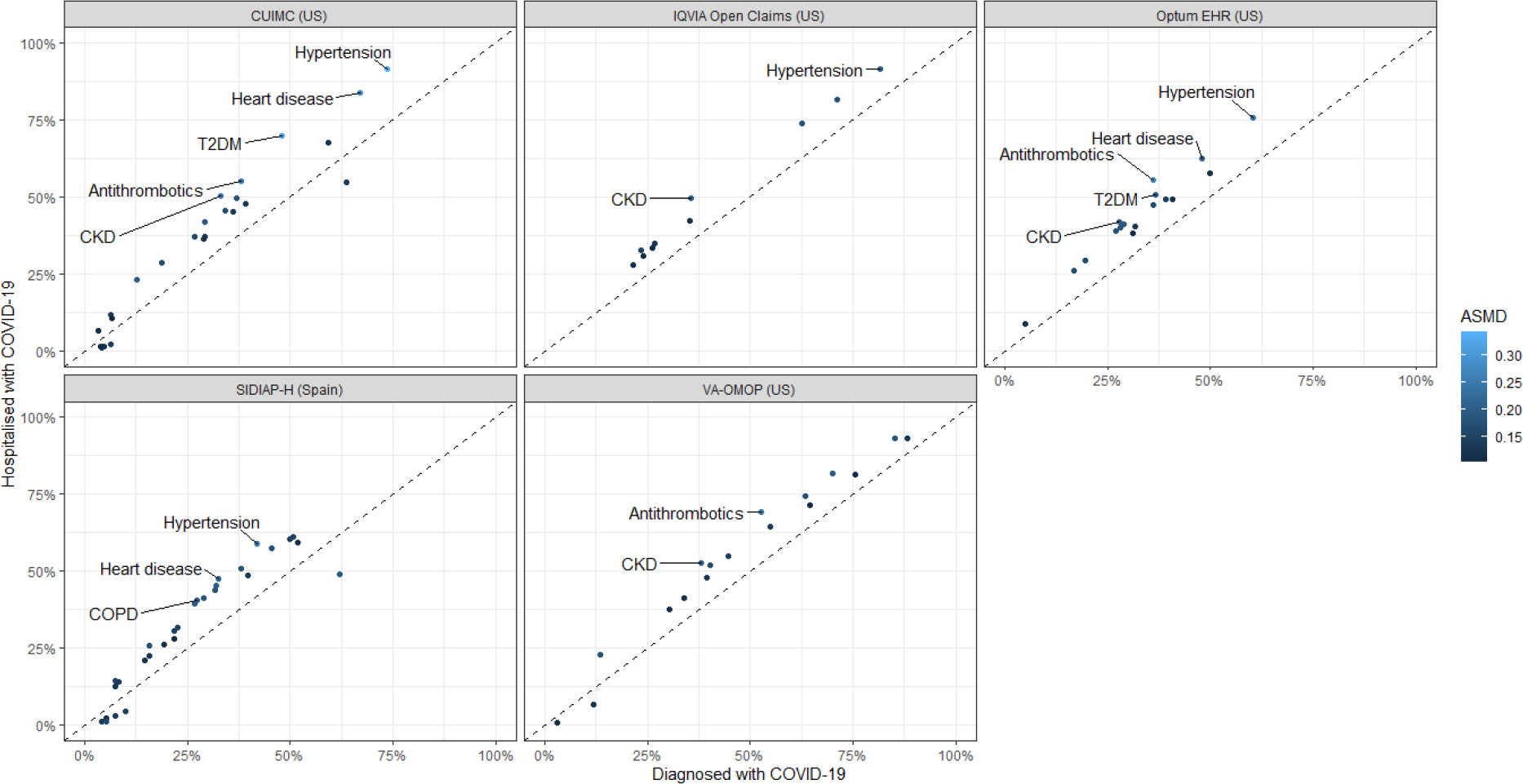
Prevalence of patient characteristics in the patients with prevalent autoimmune diseases who were diagnosed with COVID-19 compared to those hospitalised with COVID-19 This scatterplot includes patient characteristics with absolute standardised mean difference (ASMD) ≥0.1. The patient characteristics with ASMD >0.2 are labelled in the scatterplot. HIRA was not included in the scatterplot because of the significant overlap between diagnosed (n = 815) and hospitalised (n = 813) patients. CKD: chronic kidney disease; COPD: chronic obstructive pulmonary disease; T2DM: type 2 diabetes mellitus

### Study participants and follow-up

For the COVID-19 cohort, all patients diagnosed and/or hospitalised between January and June 2020 with a clinical or laboratory-confirmed diagnosis of COVID-19 and with one or more prevalent autoimmune diseases were included. For the influenza cohort, all patients diagnosed and/or hospitalised between September 2017 and April 2018 with a clinical or laboratory-confirmed diagnosis of influenza and with one or more prevalent autoimmune diseases were included. The index date (i.e., start time of the cohort) was the date of diagnosis or of hospital admission, respectively. All participants were required to have at least 365 days of observational data prior to the index date. Prevalent autoimmune condition was defined as patients having any of the following conditions captured in the data source, any time prior to the index date: Type 1 diabetes mellitus, rheumatoid arthritis, psoriasis, psoriatic arthritis, multiple sclerosis, systemic lupus erythematosus, Addison’s disease, Graves’ disease, Sjogren’s syndrome, Hashimoto thyroiditis, myasthenia gravis, vasculitis, pernicious anaemia, coeliac disease, scleroderma, sarcoidosis, ulcerative colitis, or Crohn’s disease.

Participants were followed up for the identification of study outcomes from the index date until the earliest of death, end of the study (June 2020), 30 days after index, or last date of data availability.

### Baseline characteristics

Socio-demographics (age and sex) at index date were extracted, together with comorbidities and medicines used as recorded in the 365 days prior to the index date. All features recorded in the analyzed databases were extracted, and are fully reported together with study outcomes (see below) in an aggregated form in an interactive web application (https://data.ohdsi.org/Covid19CharacterizationCharybdis/).

### Study outcomes

For the diagnosed patients, we identified hospitalisation episodes in the 30 days after the index date. For the hospitalised patients, we identified the following outcomes in the 30 days after the index date: acute myocardial infarction, cardiac arrhythmia, heart failure, stroke, venous thromboembolism, sepsis, acute respiratory distress syndrome [ARDS], pneumonia, acute kidney injury, and mortality. The outcomes were defined using code sets based on Systematized Nomenclature of Medicine (SNOMED), Current Procedural Terminology, 4th Edition (CPT), or International Classification of Diseases 9^th^ edition (ICD-9)/10^th^ edition (ICD-10) disease or procedure codes. Outcomes were not reported for the SIDIAP-H database as these were all hospital-based diagnoses and therefore highly incomplete in primary care EHR data. Mortality will only be reported for the following data sources which have good quality and complete data: CUIMC, HIRA, SIDIAP-H, and VA-OMOP.

#### Data characterisation and analysis

A common analytical package was developed based on the Observational Health Data Sciences and Informatics (OHDSI) Methods library (available at https://github.com/ohdsi-studies/Covid19CharacterizationCharybdis) and run locally in each database in a distributed network fashion. (24, 25) Results were extracted on 3^rd^ October 2020, and are constantly updated with new data in the web application.

We reported patient socio-demographics, comorbidities, and commonly used medications in the 365 days before index date. The index date for the diagnosed cohort is the earlier of the date of clinical diagnosis or laboratory confirmed diagnosis using SARS-COV2 test; whereas the index date for the hospitalised cohort is the date of admission. We calculated the absolute standardised mean difference (ASMD) for patient characteristics between the diagnosed and hospitalised with COVID-19 cohorts. We calculated the proportion of hospitalisation among diagnosed patients and the proportion of hospitalised patients having severe outcomes (acute myocardial infarction, cardiac arrhythmia, heart failure, stroke, venous thromboembolism, sepsis, ARDS, pneumonia, acute kidney injury, and mortality) within 30 days post index date.

We compared outcomes and mortality to patients with a history of autoimmune diseases hospitalised with influenza in the previous 2017-2018 season. This study was descriptive in nature, and no causal inference was intended. Multivariable regression or adjustment for confounding was therefore considered beyond the purpose and scope of our study, and not included in our study protocol. All analyses were performed and visualised using R (version 4.0.2). (26)

#### Patient and public involvement statement

No patients or public were involved in the design, execution, or dissemination of this study.

## Results

We included 133,589 patients (129,221 from US, 3,553 from Spain, and 815 from South Korea) with prevalent autoimmune diseases and a clinical diagnosis of COVID-19 or a positive SARS-CoV-2 test (Table 1). Patients were mainly female in CUIMC (63.8%), HIRA (63.4%), IQVIA Open Claims (60.5%), Optum EHR (65.9%) and SIDIAP-H (62.0%) but were predominantly male in VA-OMOP (88.3%), as expected given the population based on military veterans. The majority of cases were aged ≥50 years. Among these patients with autoimmune diseases who developed COVID-19, the most prevalent autoimmune conditions were psoriasis (3.5 to 32.5%), rheumatoid arthritis (3.9 to 18.9%), and vasculitis (3.3 to 17.5%). The most prevalent comorbidities were hypertension (25.4 to 85.2%), heart disease (32.5 to 71.1%), type 2 diabetes (21.7 to 63.3%), and hyperlipidaemia (22.7 to 59.2%). Except for HIRA, in which obesity recording rate is low, obesity was a frequently diagnosed comorbidity in all other databases (44.4 to 63.1%). The most frequently prescribed medications in the year prior to COVID-19 diagnosis across all databases were systemic antibiotics (47.2 to 84.2%), drugs used for gastroesophageal reflux disease (GERD) (39.1 to 80.6%), and non-steroidal anti-inflammatory drugs (NSAID) (31.3 to 77.5%).

**Table 1.** Baseline characteristics of study participants diagnosed with COVID-19 and had prevalent autoimmune diseases, stratified by data source

| Covariate | CUIMC<br>(US)<br>(n = 1363) | HIRA<br>(South<br>Korea)<br>(n = 815) | IQVIA<br>Open<br>Claims<br>(US)<br>(n = 104874) | Optum<br>EHR (US)<br>(n = 12897) | SIDIAP-H (Spain)<br>(n = 3553) | VA-OMOP<br>(US)<br>(n = 10087) |
| --- | --- | --- | --- | --- | --- | --- |
| **Male** | 36.2 | 36.6 | 39.5 | 34.1 | 38.0 | 88.3 |
| **Female** | 63.8 | 63.4 | 60.5 | 65.9 | 62.0 | 11.7 |
| *Age group* |  |  |  |  |  |  |
| **00-04** | <0.4 | <0.6 | 0.0 | 0.1 | <0.1 | 0.1 |
| **05-09** | <0.4 | 0.0 | 0.2 | 0.2 | 0.2 | 0.0 |
| **10-14** | <0.4 | <0.6 | 0.2 | 0.3 | 0.4 | 0.0 |
| **15-19** | 0.7 | 0.7 | 0.4 | 1.1 | 0.7 | 0.0 |
| **20-24** | 0.7 | 4.7 | 0.9 | 2.5 | 1.2 | 0.1 |
| **25-29** | 2.3 | 4.0 | 1.7 | 3.5 | 2.6 | 0.5 |
| **30-34** | 4.1 | 1.8 | 2.4 | 4.6 | 4.1 | 1.6 |
| **35-39** | 4.6 | 2.7 | 3.2 | 5.2 | 5.3 | 2.9 |
| **40-44** | 6.3 | 3.3 | 4.2 | 6.8 | 7.5 | 3.0 |
| **45-49** | 5.6 | 6.6 | 5.7 | 8.0 | 9.8 | 4.0 |
| **50-54** | 8.1 | 13.6 | 7.9 | 10.0 | 7.9 | 6.6 |
| **55-59** | 8.6 | 13.0 | 10.0 | 11.9 | 8.9 | 9.1 |
| **60-64** | 11.2 | 13.7 | 11.4 | 12.0 | 7.8 | 12.1 |
| **65-69** | 10.2 | 8.1 | 10.7 | 9.9 | 5.9 | 14.1 |
| **70-74** | 10.9 | 7.6 | 10.6 | 7.6 | 7.3 | 22.8 |
| **75-79** | 7.0 | 7.9 | 9.3 | 6.2 | 7.5 | 11.4 |
| **80-84** | 8.1 | 5.2 | 8.0 | 4.2 | 8.2 | 4.5 |
| **85-89** | 6.2 | 4.7 | 13.2 | 5.9 | 8.1 | 4.1 |
| **90-94** | 3.2 | 1.8 | 0.0 | 0.0 | 4.8 | 2.3 |
| **95-99** | 1.1 | <0.6 | 0.0 | 0.0 | 1.5 | 0.9 |
| *Autoimmune disease in the year prior to index date\** |  |  |  |  |  |  |
| **Type 1 diabetes mellitus** | 3.4 | 1.5 | 5.8 | 6.0 | 5.0 | 4.4 |
| **Rheumatoid arthritis** | 4.0 | 18.9 | 4.8 | 8.7 | 4.1 | 4.7 |
| **Psoriasis** | 3.7 | 8.2 | 3.5 | 7.4 | 27.9 | 7.1 |
| **Psoriatic arthritis** | 0.8 | 0.7 | 0.8 | 2.4 | 2.2 | 1.5 |
| **Multiple sclerosis** | 2.1 | <0.6 | 2.2 | 3.3 | 2.2 | 1.9 |
| **Systemic lupus erythematosus** | 3.4 | 1.7 | 1.9 | 3.6 | 2.3 | 1.1 |
| **Graves' disease** | 0.0 | 0.0 | 0.0 | 0.1 | 0.0 | 0.0 |
| **Hashimoto thyroiditis** | 0.0 | 0.0 | 0.0 | 2.1 | 0.0 | 0.0 |
| **Myasthenia gravis** | 0.5 | <0.6 | 0.4 | 0.6 | 1.0 | 0.6 |
| **Vasculitis** | 4.2 | 14.4 | 4.0 | 5.7 | 17.5 | 3.3 |
| **Pernicious anaemia** | 0.0 | 0.0 | 0.0 | 0.5 | 0.0 | 0.0 |
| **Coeliac disease** | 0.9 | <0.6 | 0.5 | 1.6 | 5.1 | 0.7 |
| **Scleroderma** | 0.6 | <0.6 | 0.2 | 0.4 | 0.8 | 0.2 |
| **Sarcoidosis** | 2.7 | 0.0 | 1.0 | 2.1 | 1.1 | 2.6 |
| **Ulcerative colitis** | 1.9 | <0.6 | 1.3 | 2.5 | 4.1 | 2.6 |
| **Crohn's disease** | 2.3 | <0.6 | 1.2 | 3.0 | 2.9 | 1.6 |
| *Comorbidities in the year prior to index date* |  |  |  |  |  |  |
| **Hyperlipidaemia** | 34.2 | 56.4 | 40.1 | 40.9 | 22.7 | 59.2 |
| **Asthma** | 27.0 | 28.0 | 23.3 | 22.7 | 8.3 | 15.1 |
| **CKD** | 33.0 | 14.2 | 35.6 | 27.8 | 15.8 | 38.1 |
| **COPD** | 18.8 | 3.4 | 24.1 | 16.7 | 27.4 | 40.2 |
| **Dementia** | 12.6 | 12.3 | 19.0 | 5.6 | 7.3 | 13.4 |
| **Heart disease** | 66.9 | 29.2 | 71.1 | 48.0 | 32.5 | 70.0 |
| **HIV** | 3.1 | NA | 2.1 | 0.7 | 0.4 | 2.0 |
| **Hypertension** | 73.5 | 45.6 | 81.7 | 60.4 | 42.0 | 85.2 |
| **Cancer** | 32.4 | 8.5 | 24.9 | 25.2 | 15.6 | 32.3 |
| **Obesity** | 59.4 | NA | 44.4 | 63.1 | 45.5 | 63.0 |
| **Type 2 diabetes mellitus** | 48.1 | 46.3 | 62.6 | 36.7 | 21.7 | 63.3 |
| **Cerebrovascular disease** | 6.7 | 7.7 | 8.4 | 4.7 | 3.3 | 6.9 |
| **Chronic liver disease** | 2.8 | 10.2 | 2.0 | 2.5 | 2.7 | 6.4 |
| **Pregnancy** | 2.7 | 2.0 | 1.1 | 2.3 | 0.9 | 0.1 |
| **Venous thromboembolism** | 7.4 | 3.4 | 6.1 | 4.9 | 11.5 | 6.4 |
| *Drug utilisation in the year prior to index date* |  |  |  |  |  |  |
| **Agents acting on the renin-angiotensin system** | 29.3 | 29.6 | 30.6 | 31.1 | 31.6 | 47.7 |
| **Antibacterials** | 48.2 | 84.2 | 54.2 | 49.8 | 51.7 | 54.8 |
| **Antidepressants** | 23.3 | 21.2 | 23.7 | 31.4 | 30.6 | 45.8 |
| **Antineoplastic and immunomodulating agents** | 22.9 | 10.2 | 15.6 | 21.8 | 13.3 | 21.3 |
| **Antithrombotics** | 38.0 | 47.1 | 23.5 | 36.2 | 31.9 | 52.7 |
| **Beta blockers** | 29.2 | 17.3 | 26.8 | 28.0 | 19.2 | 44.7 |
| **Calcium channel blockers** | 26.7 | 25.5 | 21.4 | 19.6 | 14.5 | 33.8 |
| **Corticosteroids** | 36.4 | 72.3 | 38.4 | 45.0 | 39.6 | 43.9 |
| **Diuretics** | 29.3 | 17.5 | 26.2 | 27.1 | 29.0 | 39.3 |
| **Drugs for obstructive airway diseases** | 31.0 | 30.2 | 39.1 | 41.2 | 28.8 | 55.1 |
| **GERD** | 39.1 | 80.6 | 51.4 | 39.2 | 50.7 | 72.8 |
| **PPI** | 28.9 | 50.2 | 24.4 | 31.8 | 49.9 | 44.6 |
| **Lipid modifying agents** | 37.0 | 34.5 | 35.2 | 36.1 | 26.8 | 64.4 |
| **NSAID** | 31.3 | 77.5 | 51.2 | 35.4 | 36.8 | 75.5 |
| **Opioids** | 24.5 | 82.1 | 24.4 | 29.0 | 21.8 | 30.4 |

Figures are presented in percentages; the figures preceded with < denote less than 5 people in that category.
\*These are not mutually exclusive, and classification is based on recent (1 year prior) records
CUIMC: Columbia University Irving Medical Center; HIRA: Health Insurance Review & Assessment Service; SIDIAP-H: Information System for Research in Primary Care – Hospitalisation Linked Data; VA-OMOP: Department of Veterans Affairs CKD: chronic kidney disease; COPD: chronic obstructive pulmonary disease; HIV: human immunodeficiency virus; GERD: gastroesophageal reflux disease; NA: Information not available; NSAID: non-steroidal anti-inflammatory drug; PPI: proton pump inhibitor

A total of 48,418 patients (46,721 from US, 884 from Spain, and 813 from South Korea) with autoimmune diseases were hospitalised with COVID-19 (Table 2). Patients were mainly female in CUIMC (54.8%), HIRA (63.5%), IQVIA Open Claims (54.8%), Optum EHR (59.5%), about equal proportion in SIDIAP-H (49.0%), but were predominantly male in VA-OMOP (93.2%). Majority of cases were aged ≥50 years. Among these patients with autoimmune diseases who were hospitalised with COVID-19, Type 1 diabetes was the most common autoimmune condition in the US databases (4.8 to 7.5%) whereas rheumatoid arthritis was most prevalent in HIRA (18.9%) and psoriasis in SIDIAP-H (30.7%). The most prevalent comorbidities were hypertension (36.5 to 93.2%), heart disease (29.0 to 83.8%), type 2 diabetes (22.4 to 74.3%), and hyperlipidaemia (27.2 to 64.5%). The most frequently prescribed medications in the year prior to hospitalisation across all databases were systemic antibiotics (52.4 to 84.0%), drugs used for gastroesophageal reflux disease (GERD) (47.9 to 80.6%), and non-steroidal anti-inflammatory drugs (NSAID) (33.0 to 81.5%). The list of patient characteristics is presented in Tables 1 and 2. A full list of the conditions that make up the prevalent autoimmune diseases is presented in Supplementary Tables 1 and 2. A complete list of patient characteristics can be found in the aforementioned interactive web application.

**Table 2.** Baseline characteristics of study participants hospitalised with COVID-19 and had prevalent autoimmune disease, stratified by data source

| Covariate | CUIMC<br>(US)<br>(n = 557) | HIRA<br>(South<br>Korea)<br>(n = 813) | IQVIA<br>Open<br>Claims<br>(US)<br>(n = 39900) | Optum<br>EHR (US)<br>(n = 3112) | SIDIAP-H (Spain)<br>(n = 884) | VA-OMOP<br>(US)<br>(n = 3152) |
| --- | --- | --- | --- | --- | --- | --- |
| **Male** | 45.2 | 36.5 | 45.2 | 40.5 | 51.0 | 93.2 |
| **Female** | 54.8 | 63.5 | 54.8 | 59.5 | 49.0 | 6.8 |
| *Age group* |  |  |  |  |  |  |
| **00-04** | <0.9 | <0.6 | 0.1 | <0.2 | <0.6 | 0.2 |
| **05-09** | <0.9 | 0.0 | 0.2 | <0.2 | 0.0 | 0.0 |
| **10-14** | <0.9 | <0.6 | 0.2 | 0.3 | 0.0 | <0.2 |
| **15-19** | <0.9 | 0.7 | 0.3 | 0.5 | <0.6 | 0.0 |
| **20-24** | <0.9 | 4.7 | 0.5 | 1.4 | <0.6 | <0.2 |
| **25-29** | 2.0 | 4.1 | 0.7 | 1.6 | 0.8 | 0.2 |
| **30-34** | 1.3 | 1.8 | 1.2 | 2.8 | 1.1 | 0.5 |
| **35-39** | 1.6 | 2.7 | 1.7 | 4.1 | 2.3 | 0.9 |
| **40-44** | 2.3 | 3.3 | 2.4 | 4.0 | 3.1 | 1.3 |
| **45-49** | 2.9 | 6.6 | 3.6 | 6.5 | 4.6 | 2.0 |
| **50-54** | 4.8 | 13.7 | 5.7 | 7.0 | 6.1 | 4.1 |
| **55-59** | 5.9 | 13.0 | 8.7 | 11.1 | 6.8 | 6.8 |
| **60-64** | 9.0 | 13.8 | 11.5 | 12.8 | 7.7 | 11.0 |
| **65-69** | 11.5 | 8.1 | 12.3 | 12.2 | 8.4 | 14.3 |
| **70-74** | 13.3 | 7.5 | 13.4 | 10.3 | 12.7 | 24.8 |
| **75-79** | 10.1 | 7.7 | 12.1 | 9.0 | 14.3 | 14.5 |
| **80-84** | 12.2 | 5.2 | 10.5 | 7.3 | 14.0 | 6.8 |
| **85-89** | 11.7 | 4.7 | 14.9 | 8.9 | 11.3 | 6.6 |
| **90-94** | 6.5 | 1.8 | 0.0 | 0.0 | 4.8 | 4.2 |
| **95-99** | 2.0 | <0.6 | 0.0 | 0.0 | 1.4 | 1.8 |
| *Autoimmune disease in the year prior to index date\** |  |  |  |  |  |  |
| **Type 1 diabetes mellitus** | 4.8 | 1.5 | 7.5 | 7.5 | 4.4 | 5.3 |
| **Rheumatoid arthritis** | 4.8 | 18.9 | 4.9 | 8.8 | 5.4 | 4.0 |
| **Psoriasis** | 1.4 | 8.2 | 2.7 | 5.4 | 26.4 | 4.4 |
| **Psoriatic arthritis** | 0.0 | 0.7 | 0.6 | 1.7 | 2.5 | 0.9 |
| **Multiple sclerosis** | 1.1 | <0.6 | 2.1 | 3.7 | 2.1 | 1.6 |
| **Systemic lupus erythematosus** | 3.2 | 1.7 | 1.9 | 4.3 | 2.6 | 0.9 |
| **Hashimoto thyroiditis** | 0.0 | 0.0 | 0.0 | 0.9 | 0.0 | 0.0 |
| **Myasthenia gravis** | <0.9 | <0.6 | 0.5 | 0.8 | 1.5 | 0.7 |
| **Vasculitis** | 3.4 | 14.4 | 4.4 | 7.7 | 20.8 | 4.4 |
| **Pernicious anaemia** | 0.0 | 0.0 | 0.0 | 0.4 | 0.0 | 0.0 |
| **Coeliac disease** | <0.9 | <0.6 | 0.3 | 0.9 | 1.2 | 0.4 |
| **Scleroderma** | <0.9 | <0.6 | 0.2 | 0.4 | 0.9 | <0.2 |
| **Sarcoidosis** | 3.4 | 0.0 | 1.2 | 1.9 | 1.2 | 2.1 |
| **Ulcerative colitis** | <0.9 | <0.6 | 1.3 | 2.2 | 2.8 | 1.6 |
| **Crohn's disease** | 1.1 | <0.6 | 1.0 | 2.4 | 2.4 | 1.2 |
| *Comorbidities in the year prior to index date* |  |  |  |  |  |  |
| **Hyperlipidaemia** | 45.8 | 56.5 | 44.9 | 49.2 | 31.9 | 64.5 |
| **Asthma** | 29.3 | 28.0 | 22.1 | 20.4 | 7.8 | 12.5 |
| **CKD** | 50.4 | 14.0 | 49.6 | 42.0 | 25.8 | 52.7 |
| **COPD** | 28.7 | 3.4 | 30.8 | 26.3 | 40.7 | 51.9 |
| **Dementia** | 23.2 | 12.2 | 21.5 | 8.9 | 6.8 | 23.1 |
| **Heart disease** | 83.8 | 29.0 | 81.7 | 62.7 | 47.7 | 81.9 |
| **HIV** | 2.7 | NA | 2.4 | 1.1 | 0.7 | 2.2 |
| **Hypertension** | 91.4 | 45.5 | 91.6 | 75.7 | 58.9 | 93.2 |
| **Cancer** | 38.4 | 8.5 | 29.0 | 29.4 | 22.6 | 37.9 |
| **Obesity** | 67.7 | NA | 48.0 | 67.0 | 57.5 | 64.0 |
| **Type 2 diabetes mellitus** | 70.0 | 46.1 | 74.1 | 50.6 | 30.8 | 74.3 |
| **Cerebrovascular disease** | 10.8 | 7.6 | 11.2 | 6.8 | 4.1 | 9.5 |
| **Chronic liver disease** | 4.1 | 10.2 | 2.9 | 3.4 | 3.3 | 9.7 |
| **Pregnancy** | 2.0 | 2.0 | 0.7 | 2.7 | 0.6 | NA |
| **Venous thromboembolism** | 11.3 | 3.4 | 8.7 | 8.8 | 14.4 | 9.3 |
| *Drug utilisation in the year prior to index date* |  |  |  |  |  |  |
| **Agents acting on the renin-angiotensin system** | 37.0 | 29.4 | 36.7 | 38.3 | 43.9 | 51.8 |
| **Antibacterials** | 52.4 | 84.0 | 55.0 | 57.6 | 59.4 | 64.4 |
| **Antidepressants** | 23.7 | 21.2 | 25.0 | 33.6 | 31.3 | 47.0 |
| **Antineoplastic and immunomodulating agents** | 23.0 | 10.0 | 15.4 | 23.3 | 18.3 | 19.3 |
| **Antithrombotics** | 55.3 | 46.9 | 32.8 | 55.5 | 45.5 | 69.3 |
| **Beta blockers** | 41.8 | 17.1 | 35.1 | 40.3 | 26.1 | 54.9 |
| **Calcium channel blockers** | 37.0 | 25.3 | 28.1 | 29.4 | 21.2 | 41.4 |
| **Corticosteroids** | 37.7 | 72.3 | 39.5 | 48.5 | 48.6 | 46.2 |
| **Diuretics** | 41.8 | 17.3 | 33.4 | 38.9 | 41.4 | 47.9 |
| **Drugs for obstructive airway diseases** | 35.7 | 30.1 | 39.8 | 46.9 | 31.9 | 58.1 |
| **GERD** | 47.9 | 80.6 | 54.1 | 49.3 | 61.2 | 77.8 |
| **PPI** | 36.6 | 50.3 | 28.0 | 40.5 | 60.3 | 48.4 |
| **Lipid modifying agents** | 49.6 | 34.6 | 42.5 | 47.4 | 39.5 | 71.4 |
| **NSAID** | 33.0 | 77.6 | 51.9 | 35.6 | 31.4 | 81.5 |
| **Opioids** | 30.7 | 82.0 | 28.0 | 41.4 | 28.2 | 37.7 |

Figures are presented in percentages; the figures preceded with < denote less than 5 people in that category
\*These are not mutually exclusive, and classification is based on recent (1 year prior) records
CUIMC: Columbia University Irving Medical Center; HIRA: Health Insurance Review & Assessment Service; SIDIAP-H: Information System for Research in Primary Care – Hospitalisation Linked Data; VA-OMOP: Department of Veterans Affairs
CKD: chronic kidney disease; COPD: chronic obstructive pulmonary disease; HIV: human immunodeficiency virus; GERD: gastroesophageal reflux disease; NA: Information not available; NSAID: non-steroidal anti-inflammatory drug; PPI: proton pump inhibitor

In patients with prevalent autoimmune diseases hospitalised with COVID-19, the prevalence of hypertension (ASMD = 0.18 to 0.34), chronic kidney disease (ASMD = 0.17 to 0.25), heart disease (ASMD = 0.18 to 0.28), Type 2 diabetes (ASMD = 0.15 to 0.32), chronic obstructive pulmonary disease (COPD) (ASMD = 0.11 to 0.20), and use of antithrombotics (ASMD = 0.15 to 0.28) were higher as compared to the larger group of such patients diagnosed with COVID-19 (Figure 1).

We included 395,784 patients with prevalent autoimmune diseases (392,797 from US, 2,419 from Spain, and 568 from South Korea) diagnosed with influenza to compare the proportion of hospitalisation episodes. The proportion of hospitalisation episodes was higher in the cohort diagnosed with COVID-19 as compared to influenza (35.7% vs 23.6% [CUIMC], 36.7% vs 16.6% [IQVIA Open Claims], 23.1% vs 17.2% [Optum EHR], 27.1% vs 19.9% [VA-OMOP], 22.2% vs 12.8% [SIDIAP-H], 96.6% vs 18.0% [HIRA]) (Figure 2).

**Figure 2.**
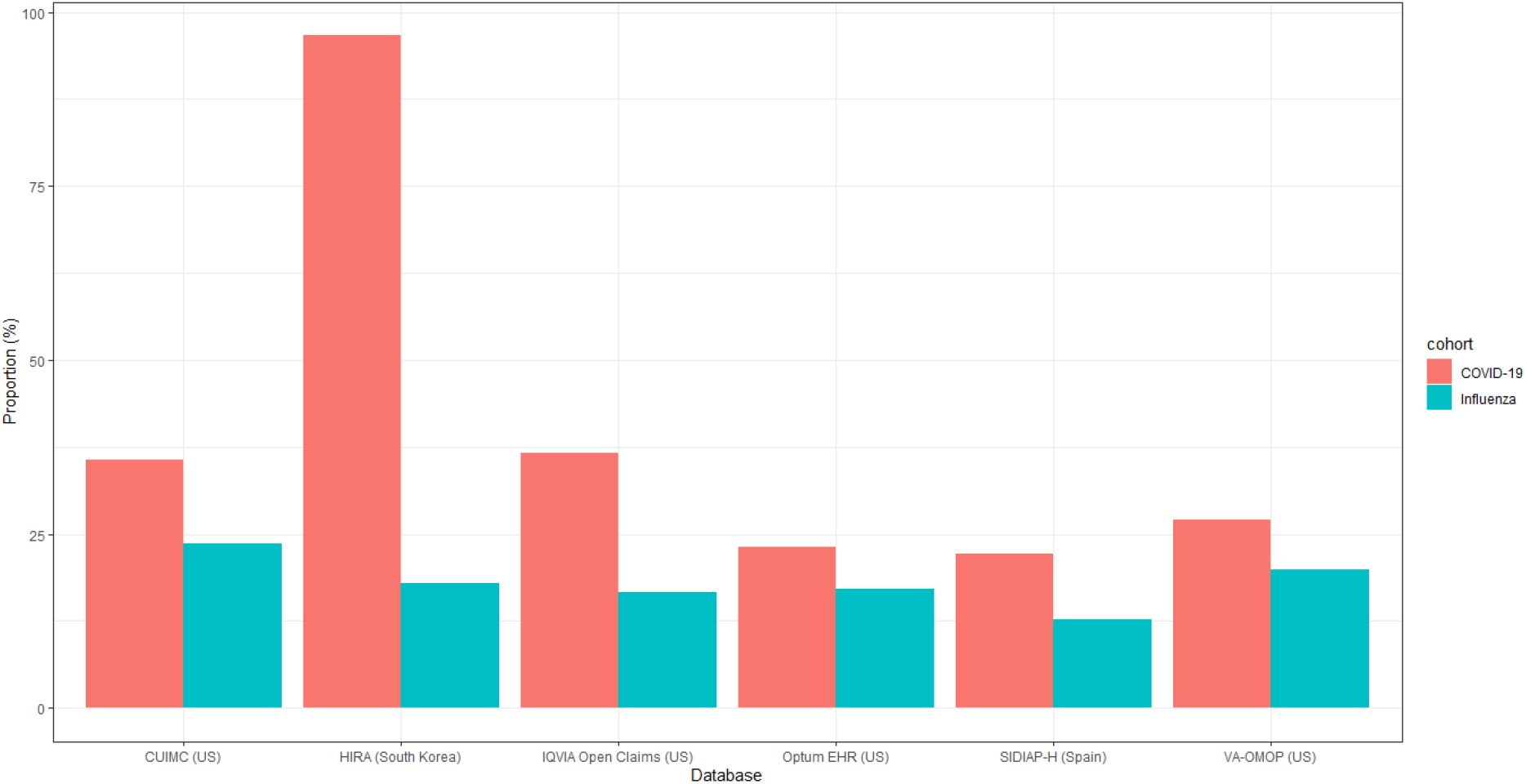
Hospitalisation in patients with prevalent autoimmune diseases in the 30-day period following a diagnosis of COVID-19 versus influenza

At 30 days post hospitalisation, the most frequent severe outcomes were related to the respiratory system, such as ARDS (2.1 to 42.8%), and pneumonia (12.6 to 53.2%) (Figure 3a). Acute kidney injury was the second most common complication, occurring in 9.9 to 31.1% of patients in US databases, and in 2.8% in HIRA. Cardiac complications were also frequent, including arrhythmia in 3.8 to 35.1% of patients, heart failure (3.9 to 24.5%), and acute myocardial infarction (2.4 to 6.3%). Sepsis occurred during hospitalisation in 4.7 to 23.5% of patients. Ischaemic or haemorrhagic stroke was recorded in 1.4 to 3.4% of patients, whereas venous thromboembolic events were recorded in 1.4 to 7.7% of patients across the databases. Mortality as a proportion of those hospitalised was generally higher in the US and Spain (16.3 to 24.6%) versus South Korea (6.3%) (Figure 3b).

**Figure 3a.**
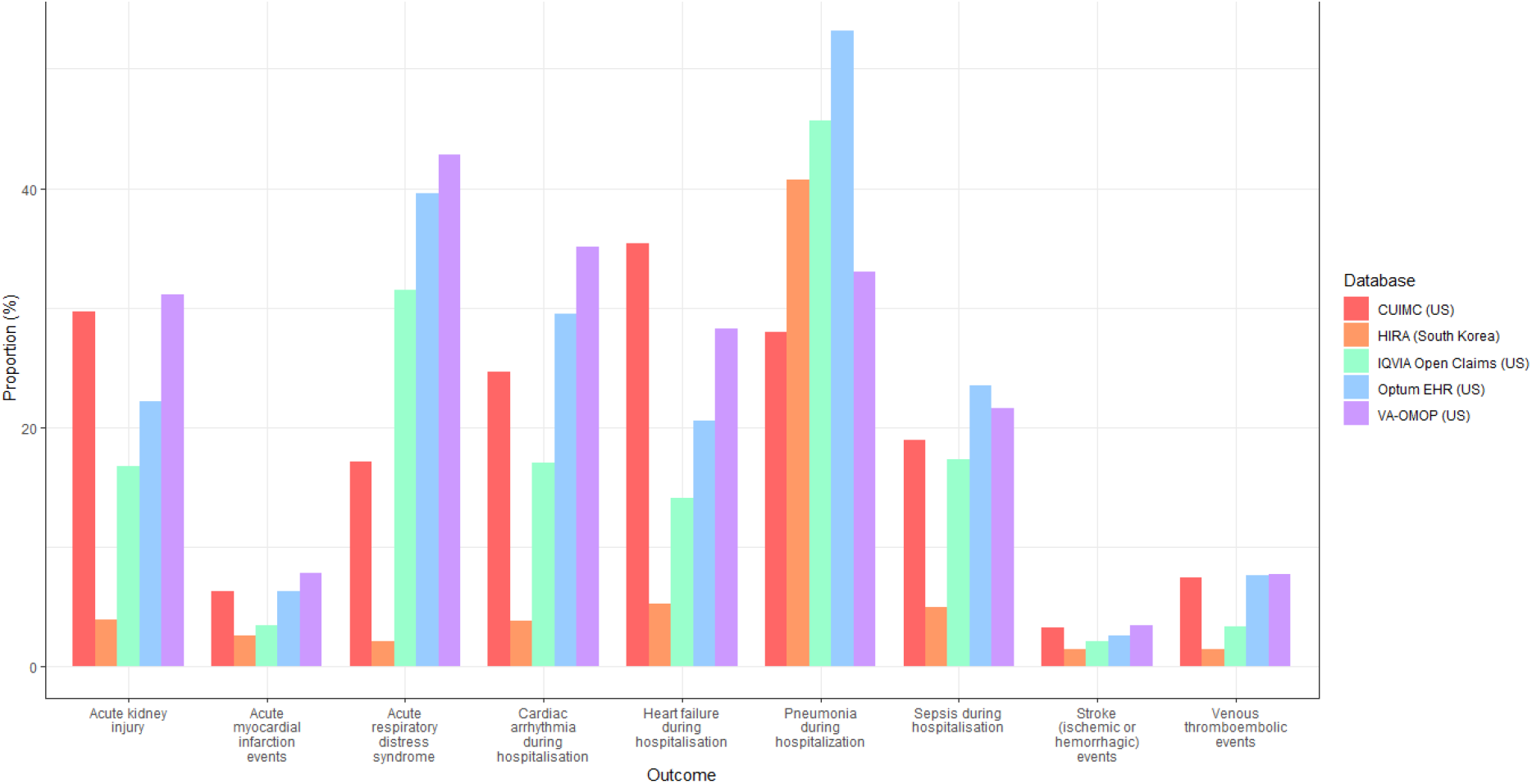
Severe outcomes in 30 days post hospital admission with COVID-19 in patients with prevalent autoimmune diseases, stratified by database

**Figure 3b.**
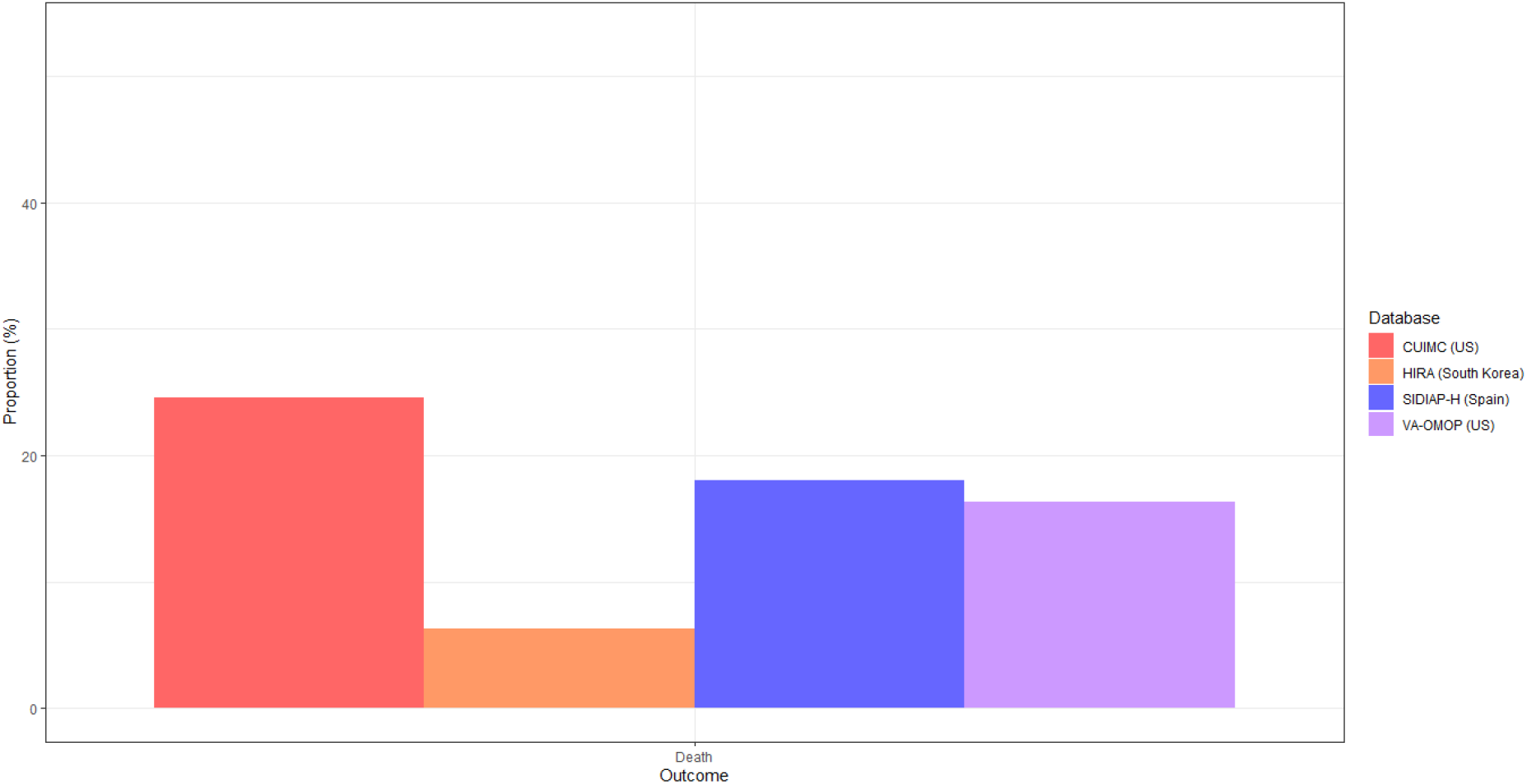
Mortality in 30 days post hospital admission with COVID-19 in patients with prevalent autoimmune diseases, stratified by database Note:Figure 3a. Hospitalisation outcomes data was not available in SIDIAP-H

Compared to 70,660 hospitalised individuals (70,184 from US, 323 from Spain, and 153 from South Korea) with influenza in previous years, patients hospitalised with COVID-19 were more likely to have higher respiratory complications such as ARDS (14.7 to 42.8% vs 16.9 to 28.7%) and pneumonia (12.6 to 53.2% vs 19.5 to 36.3%), and had a higher mortality (6.3 to 24.6% vs 2.2 to 4.4%) (Figure 4) (Supplementary table 3).

**Figure 4.**
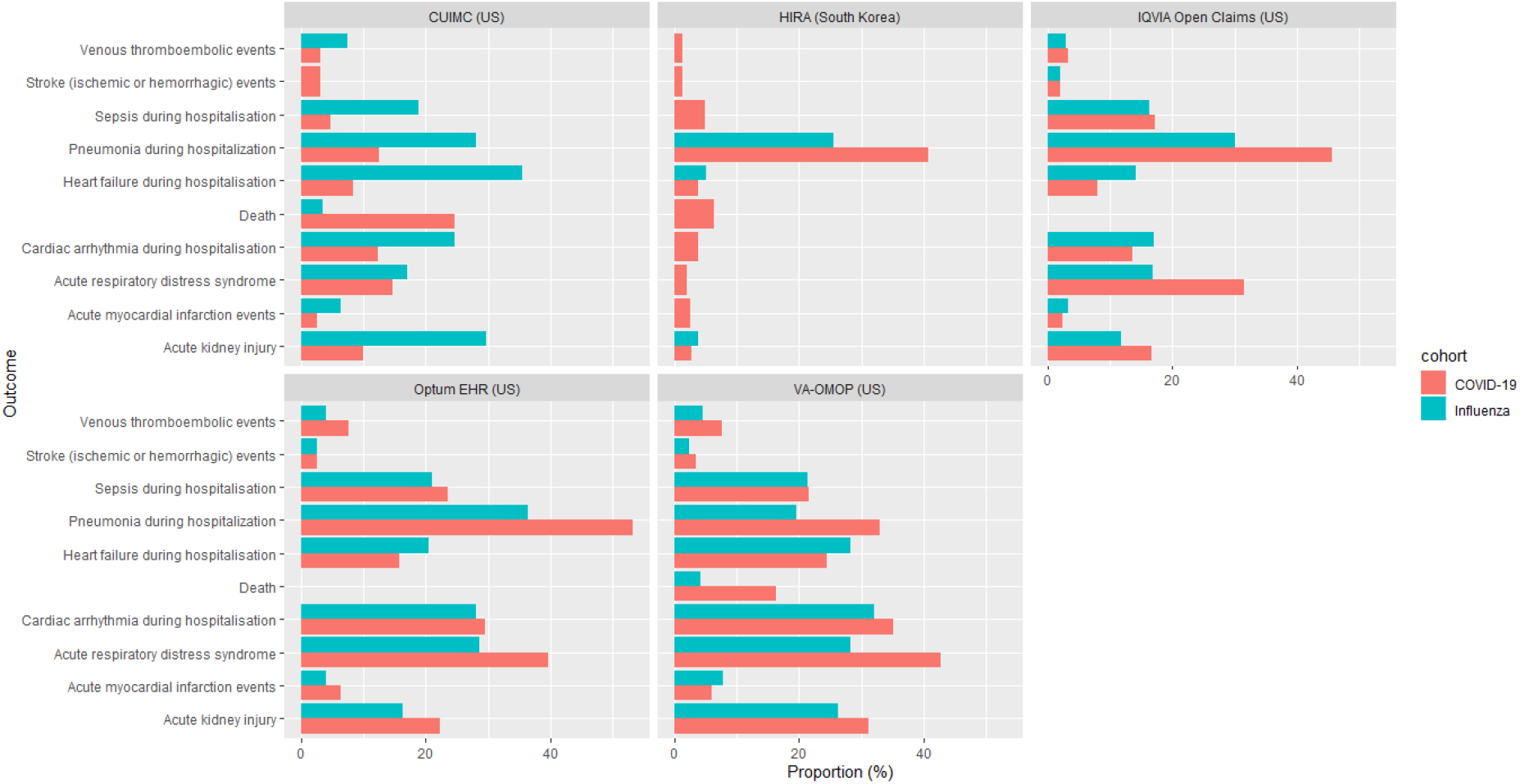
Comparison of outcomes in patients with prevalent autoimmune conditions hospitalised with COVID-19 versus influenza Note: Outcomes were omitted from the graph if there were less than 5 people experiencing the event or the data was unavailable in the respective databases.

## Discussion

This study represents the hitherto first use of routinely collected health data across the US, Spain, and South Korea to characterise hospitalised COVID-19 patients with prevalent autoimmune diseases. To our knowledge, this is the largest multinational observational study to characterise a cohort of patients with prevalent autoimmune diseases diagnosed/hospitalised with COVID-19 and detail their post-hospitalisation outcomes, during the first six months of the pandemic. We found that diagnosed autoimmune patients were predominantly female, aged above 50 years, and had pre-existing comorbidities (hypertension, heart disease, Type 2 diabetes being the most prevalent). Hospitalised autoimmune patients had similar characteristics to those diagnosed but were older and had a higher proportion of pre-existing comorbidities. As compared to patients hospitalised with influenza, more patients infected with COVID-19 died within 30 days of hospitalisation. COVID-19 patients also experienced respiratory and cardiac complications during hospitalisation.

The patients in our study were predominantly female, except for the VA-OMOP database, of which the majority were male military veterans This was consistent with the proportion of females across studies of COVID-19 in patients with autoimmune conditions in Spain (59%)(12) and in COVID-19 patients in the Global Rheumatology Alliance (GRA) physician-reported registry (67%). (19) This is likely due to females having a higher prevalence of most autoimmune diseases, but contrasts with reports of overall COVID-19 patients who were otherwise majority male. (16, 27) A recent meta-analysis has also showed that males had higher in-hospital mortality. (28) The hospitalised patients in our study were mostly aged 65 years and above, with South Korea having more patients in the age group of 50 to 64 years old. Advanced age has been reported as a poor prognostic factor for COVID-19. (15, 28) The sociodemographic profile of the patients in VA-OMOP being mostly male and older could be associated with the higher frequency of severe outcomes in that data source. The most prevalent comorbidities in our study were hypertension, heart disease, and Type 2 diabetes. This was similarly observed in the GRA registry.(19) These comorbidities were also associated with disease severity and mortality in a meta-analysis involving 12,149 general COVID-19 patients from 15 countries. (28)

Our study described post-hospitalisation complications in COVID-19 patients with prevalent autoimmune diseases. The most frequent severe outcomes in our study were ARDS, pneumonia, and cardiac injury. In the aforementioned meta-analysis (28), the most frequently reported complications associated with COVID-19 were pneumonia, respiratory failure, acute cardiac injury, and ARDS; which corroborates our findings regarding the frequency of outcomes. Cardiac injury was also independently associated with in-hospital mortality in a study conducted in Wuhan.(29) The researchers hypothesised that cardiac injury may be precipitated by acute inflammatory response as a result of COVID-infection superimposed on pre-existing cardiovascular disease. In comparison with patients hospitalised with influenza, COVID-19 patients generally had higher proportion of severe outcomes, especially respiratory complications such as ARDS and pneumonia. This phenomenon was also observed in a study conducted in a large tertiary care hospital in the US, where patients hospitalised with COVID-19 required more mechanical ventilation and had higher mortality than patients with influenza, despite presenting with less pre-existing conditions.(30) In other patient populations, such as those with obesity, patients with COVID-19 had higher mortality and requirement of intensive services as compared to similar patients with seasonal influenza, despite presenting with fewer comorbidities.(31) In pregnant women, there was a higher frequency of Caesarean section and preterm deliveries, as well as poorer outcomes (pneumonia, ARDS, sepsis, acute kidney injury, and cardiovascular and thromboembolic events) in those diagnosed with COVID-19 in comparison with seasonal influenza.(32) Like the other databases, CUIMC showed higher mortality in hospitalized patients with COVID-19 than those with influenza, but it showed lower complications in patients hospitalized with COVID-19 than influenza. A possible explanation is that patients hospitalized with influenza had higher incidence of co-morbidities like COPD and type 2 diabetes, which was also found in a previous study (33), or that data were not well captured during the height of the pandemic.

### Study limitations

COVID-19 cases may be poorly recognised due to shortages in testing capabilities, but this is to some extent mitigated in our study by also including hospitalised patients with a clinical COVID-19 diagnosis. However, even untested hospitalised patients could have been missed if hospitals were understaffed and clinicians did not have time to input proper codes. A known limitation of using routinely collected data is that medical conditions may be misclassified due to erroneous entries or underestimated as they were defined based on the presence of diagnostic or procedural codes, with the absence of records indicative of absence of disease. In particular for healthcare data in the US, the capturing of codes is largely incentivised by reimbursement from insurance companies. This factor could permit miscoding of Type 2 diabetes as Type 1 and could have enriched the autoimmune disease cohort with Type 2 diabetes patients who might not have autoimmune disease. In the initial stage of the pandemic, the lack of clinical guidance combined with the lack of access to widespread testing means that only more severe patients were seen in healthcare settings. The capture of mortality data is subject to differences by database. For example, data on inpatient deaths are recorded in a hospital EHR but deaths after discharge from hospital will not be captured in such a data source. For data sources linked to primary care, outpatient death events are typically imported into a given database from a national or local death register. It is likely that mortality rates were underestimated in our study. Nevertheless, the consistency of our findings across different healthcare settings in different countries lends credence to our results.

## Conclusions

Patients with autoimmune diseases had high rates of respiratory complications and 30-day mortality following a hospitalization with COVID-19. Compared to influenza, COVID-19 is a more severe disease, leading to more complications and higher mortality. Future studies should investigate predictors of poor outcomes in COVID-19 patients with autoimmune diseases.

## Data Availability

Analyses were performed locally in compliance with all applicable data privacy laws. Although the underlying data is not readily available to be shared, authors contributing to this paper have direct access to the data sources used in this study. All results (aggregate statistics, not presented at a patient-level with redactions for minimum cell count) are available for public inquiry. These results are inclusive of site-identifiers by contributing data sources to enable interrogation of each contributing site. All analytic code and result sets are made available at: https://github.com/ohdsi-studies/Covid19CharacterizationCharybdis

## Contributor and guarantor information

AO, FN, GH, KN, MS, KK, DPA, PBR and TDS designed the study. KK, CR, AS, SD, KL, TDS, CB, JP executed the study package on local data and contributed results. EHT, SFB analysed the data. EHT, AO, FN, and DPA interpreted the results. EHT and DPA wrote the original draft of the manuscript. All authors reviewed and edited the manuscript. The corresponding author attests that all listed authors meet authorship criteria and that no others meeting the criteria have been omitted. DPA is the guarantor.

## Copyright/license for publication

The Corresponding Author has the right to grant on behalf of all authors and does grant on behalf of all authors, a worldwide licence to the Publishers and its licensees in perpetuity, in all forms, formats and media (whether known now or created in the future), to i) publish, reproduce, distribute, display and store the Contribution, ii) translate the Contribution into other languages, create adaptations, reprints, include within collections and create summaries, extracts and/or, abstracts of the Contribution, iii) create any other derivative work(s) based on the Contribution, iv) to exploit all subsidiary rights in the Contribution, v) the inclusion of electronic links from the Contribution to third party material where-ever it may be located; and, vi) licence any third party to do any or all of the above.”

## Ethics approval

All the data partners received Institutional Review Board (IRB) approval or exemption. The use of VA data was reviewed by the Department of Veterans Affairs Central Institutional Review Board (IRB) and was determined to meet the criteria for exemption under Exemption Category 4(3) and approved the request for Waiver of HIPAA Authorization. The research was approved by the Columbia University Institutional Review Board as an OHDSI network study. The IRB number for use of HIRA data was AJIB-MED-EXP-20-065. SIDIAP analysis was approved by the Clinical Research Ethics Committee of the IDIAPJGol (project code: 20/070-PCV). Other databases used (IQVIA Open Claims and Optum EHR) are commercially available, syndicated data assets that are licensed by contributing authors for observational research. These assets are de-identified commercially available data products that could be purchased and licensed by any researcher. The collection and de-identification of these data assets is a process that is commercial intellectual property and not privileged to the data licensees and the co-authors on this study. Licensees of these data have signed Data Use Agreements with the data vendors which detail the usage protocols for running retrospective research on these databases. All analyses performed in this study were in accordance with Data Use Agreement terms as specified by the data owners. As these data are deemed commercial assets, there is no Institutional Review Board applicable to the usage and dissemination of these result sets or required registration of the protocol with additional ethics oversight. Compliance with Data Use Agreement terms, which stipulate how these data can be used and for what purpose, is sufficient for these commercial entities. Further inquiry related to the governance oversight of these assets can be made with the respective commercial entities: IQVIA (iqvia.com) and Optum (optum.com). At no point in the course of this study were the authors of this study exposed to identified patient-level data. All result sets represent aggregate, de-identified data that are represented at a minimum cell size of >5 to reduce potential for re-identification.

## Transparency declaration

Lead authors affirm that the manuscript is an honest, accurate, and transparent account of the study being reported; that no important aspects of the study have been omitted; and that any discrepancies from the study as planned have been explained.

## Appendix 1

| Data source | Source population | Sample size | Data type | Longitudinal history |
| --- | --- | --- | --- | --- |
| Columbia University Irving Medical Center (CUIMC) | Patients of the Columbia University Irving Medical Center (New York City, USA) | ≈ 6 million | The clinical data warehouse of New York-Presbyterian Hospital/Columbia University Irving Medical Center, New York, NY, based on its current and previous electronic health record systems, with data spanning over 30 years and including over 6 million patients | 1989 (1978 for diagnoses) to June 2020 |
| Health Insurance and Review Assessment (HIRA) | All citizens in South Korea | ≈ 50 million | Administrative fee-for-service claims data collected for healthcare reimbursement, including healthcare services such as treatments, pharmaceuticals, procedures, and diagnoses. | 5-years of available look-back (data older than 5-years is deleted from the database) |
| IQVIA Open Claims | USA | >300 million | A United States database of open, pre-adjudicated medical and pharmacy claims. Data are reported at anonymised patient level collected from office-based physicians and specialists via office management software and clearinghouse switch sources for the purpose of reimbursement. A subset of medical claims data have adjudicated claims. | January 2013 to May 2020 |
| Optum EHR | USA | ≈ 1.4 million | Optum® de-identified COVID-19 Electronic Health Record dataset represents Optum's Electronic Health Record data a medical records database for patients receiving a COVID-19 diagnosis record or lab test for SARS-CoV-2. The medical record data includes clinical information, inclusive of prescriptions as prescribed and administered, lab results, vital signs, body measurements, diagnoses, procedures | January 2007 to June 2020 |
| The Information System for Research in Primary Care (SIDIAP-H) | General population in Catalonia, Spain | ≈ 2 million | SIDIAP is a primary care records database that covers approximately 7 million people, equivalent to an 80% of the population of Catalonia, North-East Spain. The SIDIAP-H subset of the database includes around 2 million people out of the total 7 million in SIDIAP that are registered in primary care practices with linked hospital inpatient data (up to 2018 only) available as obtained from the Catalan Institute of Health hospitals. Healthcare is universal and tax-payer funded in the region, and primary care physicians are gatekeepers for all care and responsible for repeat prescriptions. | 2006 to June 2020 |
| United States Department of Veterans Affairs (VA-OMOP) | Patients of the Veterans Affairs in the United States | ≈ 9 million | VA OMOP data reflects the national Department of Veterans Affairs health care system, which is the largest integrated provider of medical and mental health services in the United States. Care is provided at 170 VA Medical Centers and 1,063 outpatient sites serving more than 9 million enrolled Veterans each year. | 2000 to August 2020 |

## References

1. Johns Hopkins University. COVID-19 Dashboard by the Center for Systems Science and Engineering (CSSE) at Johns Hopkins University [updated 27 August 2020. Available from: https://coronavirus.jhu.edu/map.html.

2. Kastritis E, Kitas GD, Vassilopoulos D, Giannopoulos G, Dimopoulos MA, Sfikakis PP. Systemic autoimmune diseases, anti-rheumatic therapies, COVID-19 infection risk and patient outcomes. Rheumatology International. 2020;40(9):1353–60.

3. Furman D, Campisi J, Verdin E, Carrera-Bastos P, Targ S, Franceschi C, et al. Chronic inflammation in the etiology of disease across the life span. Nat Med. 2019;25(12):1822–32.

4. Hollan I, Meroni PL, Ahearn JM, Cohen Tervaert JW, Curran S, Goodyear CS, et al. Cardiovascular disease in autoimmune rheumatic diseases. Autoimmunity Reviews. 2013;12(10):1004–15.

5. Sitia S, Atzeni F, Sarzi-Puttini P, Di Bello V, Tomasoni L, Delfino L, et al. Cardiovascular involvement in systemic autoimmune diseases. Autoimmunity Reviews. 2009;8(4):281–6.

6. Yusuf HR, Hooper WC, Beckman MG, Zhang QC, Tsai J, Ortel TL. Risk of venous thromboembolism among hospitalizations of adults with selected autoimmune diseases. J Thromb Thrombolysis. 2014;38(3):306–13.

7. Ramagopalan SV, Wotton CJ, Handel AE, Yeates D, Goldacre MJ. Risk of venous thromboembolism in people admitted to hospital with selected immune-mediated diseases: record-linkage study. BMC Medicine. 2011;9(1):1.

8. Zöller B, Li X, Sundquist J, Sundquist K. Autoimmune diseases and venous thromboembolism: a review of the literature. Am J Cardiovasc Dis. 2012;2(3):171–83.

9. Chen G, Wu D, Guo W, Cao Y, Huang D, Wang H, et al. Clinical and immunological features of severe and moderate coronavirus disease 2019. J Clin Invest. 2020;130(5):2620–9.

10. Huang C, Wang Y, Li X, Ren L, Zhao J, Hu Y, et al. Clinical features of patients infected with 2019 novel coronavirus in Wuhan, China. Lancet. 2020;395(10223):497–506.

11. Wu C, Chen X, Cai Y, Xia Ja, Zhou X, Xu S, et al. Risk Factors Associated With Acute Respiratory Distress Syndrome and Death in Patients With Coronavirus Disease 2019 Pneumonia in Wuhan, China. JAMA Internal Medicine. 2020;180(7):934–43.

12. Freites Nuñez DD, Leon L, Mucientes A, Rodriguez-Rodriguez L, Font Urgelles J, Madrid García A, et al. Risk factors for hospital admissions related to COVID-19 in patients with autoimmune inflammatory rheumatic diseases. Annals of the Rheumatic Diseases. 2020:annrheumdis-2020-217984.

13. Haberman R, Axelrad J, Chen A, Castillo R, Yan D, Izmirly P, et al. Covid-19 in Immune-Mediated Inflammatory Diseases - Case Series from New York. N Engl J Med. 2020;383(1):85–8.

14. Nicola M, O’Neill N, Sohrabi C, Khan M, Agha M, Agha R. Evidence based management guideline for the COVID-19 pandemic - Review article. Int J Surg. 2020;77:206–16.

15. Tam L-S, Tanaka Y, Handa R, Chang C-C, Cheng YK, Isalm N, et al. Care for patients with rheumatic diseases during COVID-19 pandemic: A position statement from APLAR. Int J Rheum Dis. 2020;23(6):717–22.

16. Huang Y, Chen Z, Wang Y, Han L, Qin K, Huang W, et al. Clinical characteristics of 17 patients with COVID-19 and systemic autoimmune diseases: a retrospective study. Annals of the Rheumatic Diseases. 2020;79(9):1163–9.

17. Mathian A, Mahevas M, Rohmer J, Roumier M, Cohen-Aubart F, Amador-Borrero B, et al. Clinical course of coronavirus disease 2019 (COVID-19) in a series of 17 patients with systemic lupus erythematosus under long-term treatment with hydroxychloroquine. Annals of the Rheumatic Diseases. 2020;79(6):837–9.

18. Monti S, Balduzzi S, Delvino P, Bellis E, Quadrelli VS, Montecucco C. Clinical course of COVID-19 in a series of patients with chronic arthritis treated with immunosuppressive targeted therapies. Annals of the Rheumatic Diseases. 2020;79(5):667–8.

19. Gianfrancesco M, Hyrich KL, Al-Adely S, Carmona L, Danila MI, Gossec L, et al. Characteristics associated with hospitalisation for COVID-19 in people with rheumatic disease: data from the COVID-19 Global Rheumatology Alliance physician-reported registry. Annals of the Rheumatic Diseases. 2020;79(7):859–66.

20. Anthony Sena, Kristin Kostka, Martijn Schuemie, jdposada. ohdsi-studies/Covid19CharacterizationCharybdis: Charybdis v1.1.1 - Publication Package (Version v1.1.1): Zenodo; 2020 [updated 2020, September 16. Available from: http://doi.org/10.5281/zenodo.4033034.

21. Voss EA, Makadia R, Matcho A, Ma Q, Knoll C, Schuemie M, et al. Feasibility and utility of applications of the common data model to multiple, disparate observational health databases. Journal of the American Medical Informatics Association. 2015;22(3):553–64.

22. García-Gil Mdel M, Hermosilla E, Prieto-Alhambra D, Fina F, Rosell M, Ramos R, et al. Construction and validation of a scoring system for the selection of high-quality data in a Spanish population primary care database (SIDIAP). Inform Prim Care. 2011;19(3):135–45.

23. Kim J-A, Yoon S, Kim L-Y, Kim D-S. Towards Actualizing the Value Potential of Korea Health Insurance Review and Assessment (HIRA) Data as a Resource for Health Research: Strengths, Limitations, Applications, and Strategies for Optimal Use of HIRA Data. J Korean Med Sci. 2017;32(5):718–28.

24. Hripcsak G, Duke JD, Shah NH, Reich CG, Huser V, Schuemie MJ, et al. Observational Health Data Sciences and Informatics (OHDSI): Opportunities for Observational Researchers. Stud Health Technol Inform. 2015;216:574–8.

25. Hripcsak G, Ryan PB, Duke JD, Shah NH, Park RW, Huser V, et al. Characterizing treatment pathways at scale using the OHDSI network. Proc Natl Acad Sci U S A. 2016;113(27):7329–36.

26. R Development Core Team. R: A language and environment for statistical computing. Vienna, Austria; 2013.

27. Argenziano MG, Bruce SL, Slater CL, Tiao JR, Baldwin MR, Barr RG, et al. Characterization and clinical course of 1000 patients with coronavirus disease 2019 in New York: retrospective case series. BMJ. 2020;369:m1996.

28. Jutzeler CR, Bourguignon L, Weis CV, Tong B, Wong C, Rieck B, et al. Comorbidities, clinical signs and symptoms, laboratory findings, imaging features, treatment strategies, and outcomes in adult and pediatric patients with COVID-19: A systematic review and meta-analysis. Travel Med Infect Dis. 2020;37:101825-.

29. Shi S, Qin M, Shen B, Cai Y, Liu T, Yang F, et al. Association of Cardiac Injury With Mortality in Hospitalized Patients With COVID-19 in Wuhan, China. JAMA Cardiol. 2020;5(7):802–10.

30. Donnino MW, Moskowitz A, Thompson GS, Heydrick SJ, Pawar RD, Berg KM, et al. Comparison Between Influenza and COVID-19 at a Tertiary Care Center. medRxiv. 2020:2020.08.19.20163857.

31. Recalde M, Roel E, Pistillo A, Sena AG, Prats-Uribe A, Ahmed WU-R, et al. Characteristics and outcomes of 627 044 COVID-19 patients with and without obesity in the United States, Spain, and the United Kingdom. medRxiv. 2020:2020.09.02.20185173.

32. Lai LYH, Golozar A, Sena A, Margulis AV, Haro N, Casajust P, et al. “Clinical characteristics, symptoms, management and health outcomes in 8,598 pregnant women diagnosed with COVID-19 compared to 27,510 with seasonal influenza in France, Spain and the US: a network cohort analysis”. medRxiv. 2020:2020.10.13.20211821.

33. Burn E, You SC, Sena AG, Kostka K, Abedtash H, Abrahão MTF, et al. Deep phenotyping of 34,128 adult patients hospitalised with COVID-19 in an international network study. Nature Communications. 2020;11(1):5009.

