## Supplementary files for "Characteristics, outcomes, and mortality amongst 133,589 patients with prevalent autoimmune diseases diagnosed with, and 48,418 hospitalised for COVID-19: a multinational distributed network cohort analysis"

**Supplementary Figure 1. Database selection process**

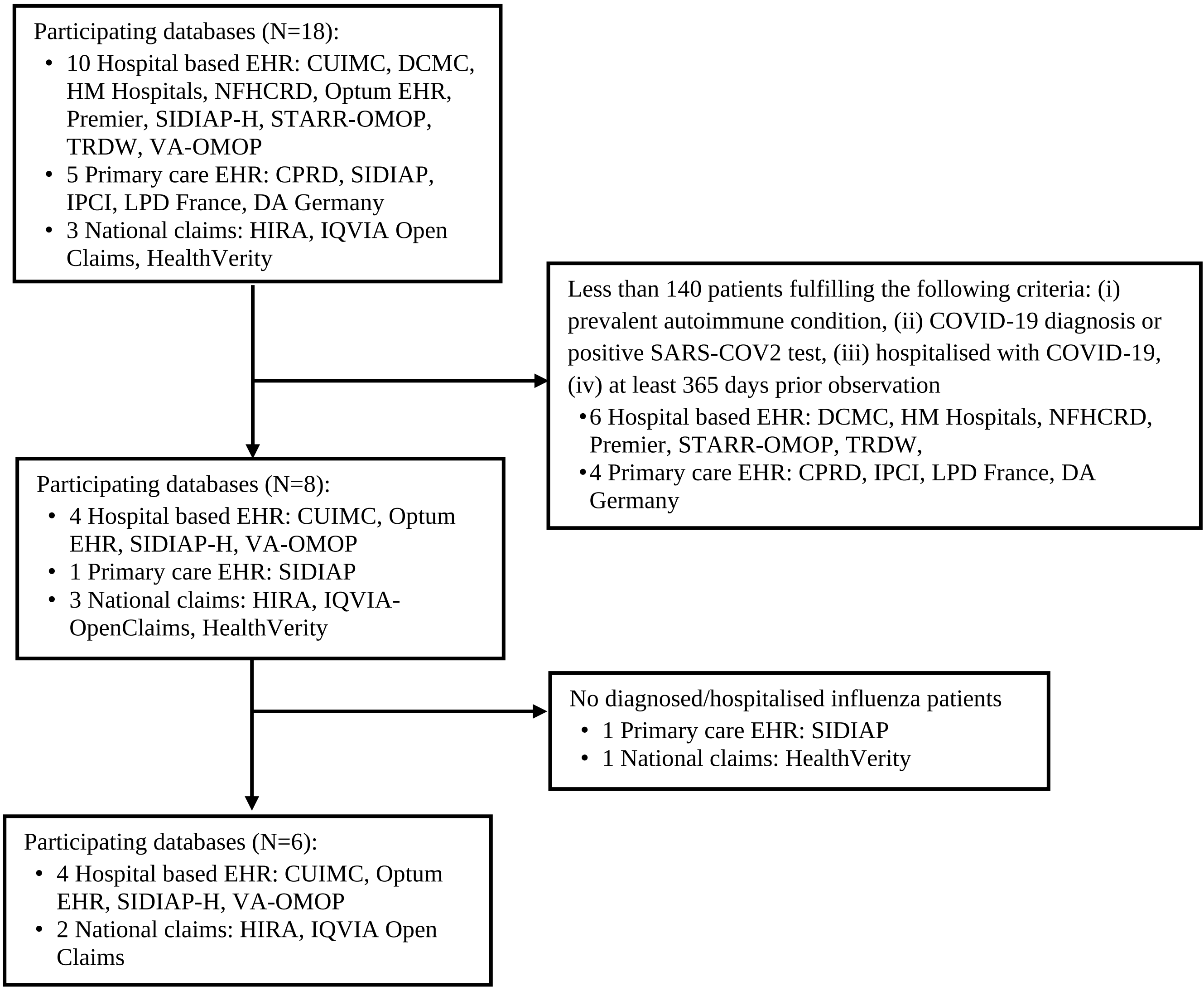


CPRD: Clinical Practice Research Datalink; CUIMC: Columbia University Irving Medical Center; DA Germany: IQVIA disease analyser Germany; DCMC: Daegu Catholic University Medical Center; EHR: Electronic health record; HIRA: Health Insurance Review & Assessment Service; LPD France: IQVIA Longitudinal Patient Data France; NFHCRD: Nanfang Hospital COVID-19 Research Database; IPCI: Integrated Primary Care Information; SIDIAP: Information System for Research in Primary Care; SIDIAP-H: SIDIAP– Hospitalisation Linked Data; TRDW: Tufts Research Data Warehouse, VA-OMOP: Department of Veterans Affairs

**Supplementary table 1. Prevalence of autoimmune diseases in the year prior to the index date in patients diagnosed with COVID-19**

The condition with the highest prevalence in most databases (highlighted in yellow) was reported as the prevalence for the respective autoimmune disease in Table 1 of the manuscript.

| Autoimmune disease | Condition | CUIMC | HIRA | IQVIA-Open Claims | Optum EHR | SIDIAP-H | VA-OMOP |
|---|---|---|---|---|---|---|---|
| **Type 1 Diabetes Mellitus** | acidosis due to type 1 diabetes mellitus | 0.0 | 0.0 | 0.0 | 0.3 | 0.0 | 0.0 |
| | disorder due to type 1 diabetes mellitus | 3.3 | 2.0 | 3.9 | 4.8 | 1.2 | 4.4 |
| | gangrene due to type 1 diabetes mellitus | 0.0 | 0.0 | 0.1 | 0.1 | 0.0 | 0.0 |
| | hyperglycemia due to type 1 diabetes mellitus | 1.9 | 0.0 | 1.9 | 2.9 | <0.1 | 1.6 |
| | hypoglycemia due to type 1 diabetes mellitus | 0.5 | 0.0 | 0.4 | 0.8 | 0.0 | 0.4 |
| | mild nonproliferative retinopathy due to type 1 diabetes mellitus | <0.4 | 0.0 | 0.1 | 0.3 | 0.0 | 0.3 |
| | moderate nonproliferative retinopathy due to type 1 diabetes mellitus | 0.0 | 0.0 | 0.1 | 0.1 | 0.0 | 0.2 |
| | neuropathic arthropathy due to type 1 diabetes mellitus | 0.0 | 0.0 | 0.0 | 0.1 | 0.0 | 0.0 |
| | neuropathy due to type 1 diabetes mellitus | 0.5 | 0.0 | 0.7 | 0.8 | <0.1 | 0.6 |
| | nonproliferative diabetic retinopathy due to type 1 diabetes mellitus | <0.4 | 0.0 | 0.2 | 0.4 | 0.0 | 0.5 |
| | peripheral circulatory disorder due to type 1 diabetes mellitus | 0.6 | <0.6 | 0.4 | 0.4 | 0.6 | 0.2 |
| | peripheral neuropathy due to type 1 diabetes mellitus | 0.4 | 0.0 | 0.5 | 0.7 | <0.1 | 0.4 |
| | polyneuropathy due to type 1 diabetes mellitus | 0.4 | 0.0 | 0.5 | 0.6 | <0.1 | 0.4 |
| | pre-existing type 1 diabetes mellitus | <0.4 | 0.0 | 0.0 | 0.1 | <0.1 | 0.0 |
| | pre-existing type 1 diabetes mellitus in pregnancy | <0.4 | 0.0 | 0.0 | 0.1 | <0.1 | 0.0 |
| | pregnancy and type 1 diabetes mellitus | <0.4 | 0.0 | 0.0 | 0.1 | <0.1 | 0.0 |
| | renal disorder due to type 1 diabetes mellitus | 1.1 | <0.6 | 0.9 | 1.2 | 0.3 | 0.6 |
| | type 1 diabetes mellitus | 3.4 | 1.5 | 5.8 | 6.0 | 5.0 | 4.4 |

| | | | | | | | |
|---|---|---|---|---|---|---|---|
| | type 1 diabetes mellitus uncontrolled | 0.0 | 0.0 | 0.0 | 0.8 | 0.0 | 0.0 |
| | type 1 diabetes mellitus with arthropathy | 0.0 | 0.0 | 0.0 | 0.1 | 0.0 | 0.0 |
| | type 1 diabetes mellitus without complication | 2.6 | 1.0 | 3.7 | 3.8 | 4.9 | 2.2 |
| | ulcer of lower limb due to type 1 diabetes mellitus | <0.4 | 0.0 | 0.2 | 0.3 | <0.1 | 0.3 |
| **Rheumatoid arthritis** | myopathy due to rheumatoid arthritis | 0.0 | 0.0 | 0.1 | 0.0 | 0.0 | 0.0 |
| | polyneuropathy in rheumatoid arthritis | 0.0 | 0.0 | 0.1 | 0.0 | 0.0 | 0.0 |
| | rheumatoid arthritis | 4.0 | 18.9 | 4.8 | 8.7 | 4.1 | 4.7 |
| | rheumatoid arthritis - ankle and/or foot | <0.4 | 0.0 | 0.1 | 0.1 | 0.0 | 0.1 |
| | rheumatoid arthritis - hand joint | 0.5 | 0.0 | 0.2 | 0.5 | <0.1 | 0.2 |
| | rheumatoid arthritis of elbow | 0.0 | 0.0 | 0.0 | 0.1 | 0.0 | 0.0 |
| | rheumatoid arthritis of knee | <0.4 | 0.0 | 0.1 | 0.2 | 0.0 | 0.1 |
| | rheumatoid arthritis of shoulder | 0.0 | 0.0 | 0.1 | 0.2 | 0.0 | 0.0 |
| | rheumatoid arthritis of wrist | 0.0 | 0.0 | 0.1 | 0.1 | 0.0 | 0.1 |
| | rheumatoid lung disease with rheumatoid arthritis | 0.0 | 0.0 | 0.1 | 0.0 | 0.0 | 0.1 |
| | seronegative rheumatoid arthritis | 1.2 | 2.3 | 0.9 | 1.7 | 0.9 | 0.6 |
| | seropositive rheumatoid arthritis | 1.9 | 5.0 | 2.0 | 3.9 | 2.9 | 1.9 |
| **Psoriasis** | arthritis mutilans | 0.0 | 0.7 | 0.1 | 0.1 | 0.0 | 0.6 |
| | generalized pustular psoriasis | 0.0 | <0.6 | 0.0 | 0.0 | <0.1 | 0.1 |
| | guttate psoriasis | <0.4 | <0.6 | 0.0 | 0.2 | 1.9 | 0.2 |
| | localized pustular psoriasis | 0.0 | <0.6 | 0.0 | 0.1 | <0.1 | 0.0 |
| | psoriasis | 3.7 | 8.2 | 3.5 | 7.4 | 27.9 | 7.1 |
| | psoriasis vulgaris | <0.4 | 2.8 | 1.3 | 1.0 | 0.3 | 2.6 |
| | psoriasis with arthropathy | 0.8 | 0.7 | 0.8 | 2.4 | 2.2 | 1.5 |
| | psoriatic arthritis | 0.0 | 0.0 | 0.0 | 0.1 | <0.1 | 0.1 |
| | psoriatic arthritis mutilans | 0.0 | 0.0 | 0.1 | 0.1 | 0.0 | 0.6 |
| | pustular psoriasis | 0.0 | 0.9 | 0.1 | 0.2 | <0.1 | 0.1 |
| | pustular psoriasis of palm of hand | 0.0 | <0.6 | 0.0 | 0.1 | <0.1 | 0.0 |
| | pustular psoriasis of palms and soles | 0.0 | <0.6 | 0.0 | 0.1 | <0.1 | 0.0 |
| | pustular psoriasis of sole of foot | 0.0 | <0.6 | 0.0 | 0.1 | <0.1 | 0.0 |

| | | | | | | | |
|---|---|---|---|---|---|---|---|
| **Psoriatic Arthritis** | arthritis mutilans | 0.0 | 0.7 | 0.1 | 0.1 | 0.0 | 0.6 |
| | psoriasis with arthropathy | 0.8 | 0.7 | 0.8 | 2.4 | 2.2 | 1.5 |
| | psoriatic arthritis mutilans | 0.0 | 0.0 | 0.1 | 0.1 | 0.0 | 0.6 |
| **Multiple sclerosis** | multiple sclerosis | 2.1 | <0.6 | 2.2 | 3.3 | 2.2 | 1.9 |
| **Systemic lupus erythematosus** | drug-induced systemic lupus erythematosus | <0.4 | 0.0 | 0.0 | 0.1 | 0.0 | 0.0 |
| | lung disease with systemic lupus erythematosus | <0.4 | 0.0 | 0.0 | 0.1 | 0.0 | 0.0 |
| | nephropathy co-occurrent and due to systemic lupus erythematosus | 0.8 | 0.0 | 0.2 | 0.6 | <0.1 | 0.1 |
| | nephrosis co-occurrent and due to systemic lupus erythematosus | 0.8 | 0.0 | 0.2 | 0.6 | <0.1 | 0.1 |
| | sle glomerulonephritis syndrome | 0.8 | 0.0 | 0.2 | 0.6 | <0.1 | 0.1 |
| | systemic lupus erythematosus | 3.4 | 1.7 | 1.9 | 3.6 | 2.3 | 1.1 |
| | systemic lupus erythematosus with organ/system involvement | 1.2 | 1.2 | 0.4 | 1.1 | <0.1 | 0.2 |
| | systemic lupus erythematosus with pericarditis | <0.4 | 0.0 | 0.0 | 0.1 | 0.0 | 0.0 |
| **Graves' disease** | graves' disease | 0.0 | 0.0 | 0.0 | 0.1 | 0.0 | 0.0 |
| **Hashimoto thyroiditis** | hashimoto thyroiditis | 0.0 | 0.0 | 0.0 | 2.1 | 0.0 | 0.0 |
| **Myasthenia gravis** | myasthenia gravis | 0.5 | <0.6 | 0.4 | 0.6 | 1.0 | 0.6 |
| | myasthenia gravis with exacerbation | <0.4 | 0.0 | 0.1 | 0.1 | <0.1 | 0.2 |
| | myasthenia gravis without exacerbation | 0.4 | 0.0 | 0.4 | 0.6 | 1.0 | 0.5 |
| **Vasculitis** | acute febrile mucocutaneous lymph node syndrome | <0.4 | <0.6 | 0.0 | 0.1 | <0.1 | 0.0 |
| | acute hemorrhagic gastritis | 0.0 | 6.9 | 0.3 | 0.3 | <0.1 | 0.3 |
| | antineutrophil cytoplasmic antibody positive vasculitis | <0.4 | <0.6 | 0.1 | 0.2 | <0.1 | 0.1 |
| | arteritis | 1.3 | <0.6 | 0.7 | 1.1 | 3.5 | 0.6 |
| | autoimmune vasculitis | <0.4 | <0.6 | 0.0 | 0.2 | <0.1 | 0.0 |
| | behcet's syndrome | <0.4 | 0.9 | 0.0 | 0.1 | 0.4 | 0.1 |
| | capillaritis | <0.4 | <0.6 | 0.1 | 0.0 | <0.1 | 0.1 |
| | deep thrombophlebitis | <0.4 | 1.6 | 0.3 | 0.1 | 6.6 | 0.2 |
| | deep vein phlebitis and thrombophlebitis of the leg | <0.4 | 1.6 | 0.3 | 0.1 | 6.6 | 0.2 |

| | | | | | | | |
|---|---|---|---|---|---|---|---|
| | giant cell arteritis with polymyalgia rheumatica | 0.0 | 0.0 | 0.0 | 0.0 | <0.1 | 0.1 |
| | granulomatosis with polyangiitis | <0.4 | 0.0 | 0.1 | 0.2 | <0.1 | 0.1 |
| | hypersensitivity angiitis | <0.4 | <0.6 | 0.1 | 0.3 | <0.1 | 0.0 |
| | idiopathic capillaritis | <0.4 | <0.6 | 0.1 | 0.0 | <0.1 | 0.1 |
| | necrotizing vasculitis | <0.4 | <0.6 | 0.1 | 0.1 | 0.8 | 0.1 |
| | phlebitis | 2.1 | 4.7 | 2.6 | 3.6 | 12.8 | 1.8 |
| | phlebitis and thrombophlebitis | 0.7 | 1.6 | 0.9 | 1.1 | 10.2 | 0.6 |
| | phlebitis and thrombophlebitis of intracranial sinuses | 0.0 | 0.0 | 0.0 | 0.1 | 0.0 | 0.0 |
| | phlebitis of deep veins of lower extremity | <0.4 | 1.6 | 0.4 | 0.2 | 6.6 | 0.3 |
| | phlebitis of lower limb vein | 1.5 | 2.9 | 2.0 | 2.6 | 9.2 | 1.3 |
| | phlebitis of superficial veins of lower extremity | <0.4 | <0.6 | 0.4 | 0.5 | 0.1 | 0.3 |
| | phlebitis of the femoral vein | 0.0 | 0.0 | 0.1 | 0.0 | <0.1 | 0.1 |
| | pigmented purpuric lichenoid dermatitis of gougerot and blum | <0.4 | <0.6 | 0.1 | 0.0 | <0.1 | 0.1 |
| | polyarteritis | 0.0 | <0.6 | 0.1 | 0.1 | <0.1 | 0.1 |
| | polyarteritis nodosa | 0.0 | <0.6 | 0.1 | 0.1 | <0.1 | 0.1 |
| | primary systemic arteritis | <0.4 | <0.6 | 0.1 | 0.1 | 0.2 | 0.1 |
| | primary systemic vasculitis | 0.5 | 1.7 | 0.2 | 0.5 | 0.6 | 0.3 |
| | retinal vasculitis | <0.4 | 0.0 | 0.0 | 0.0 | 0.0 | 0.1 |
| | rheumatoid vasculitis | <0.4 | <0.6 | 0.0 | 0.1 | <0.1 | 0.0 |
| | secondary systemic vasculitis | <0.4 | <0.6 | 0.1 | 0.2 | <0.1 | 0.1 |
| | small vessel vasculitis | <0.4 | <0.6 | 0.2 | 0.4 | 0.2 | 0.2 |
| | small vessel vasculitis caused by immune complex | <0.4 | <0.6 | 0.0 | 0.2 | <0.1 | 0.0 |
| | systemic vasculitis | 0.9 | 2.6 | 0.4 | 0.7 | 0.8 | 0.4 |
| | temporal arteritis | <0.4 | <0.6 | 0.3 | 0.4 | 0.8 | 0.3 |
| | thromboangiitis | <0.4 | <0.6 | 0.0 | 0.0 | <0.1 | 0.1 |
| | thromboangiitis obliterans | <0.4 | <0.6 | 0.0 | 0.0 | <0.1 | 0.1 |
| | thrombophlebitis | 0.7 | 3.3 | 0.9 | 1.2 | 10.2 | 0.6 |
| | thrombophlebitis of deep veins of lower extremity | <0.4 | 1.6 | 0.3 | 0.1 | 6.6 | 0.2 |

| | | | | | | | |
|---|---|---|---|---|---|---|---|
| | thrombophlebitis of lower extremities | <0.4 | 2.3 | 0.3 | 0.1 | 6.6 | 0.2 |
| | varicose veins of lower extremity with inflammation | 1.2 | 1.2 | 1.4 | 1.9 | 2.5 | 0.8 |
| | varicose veins of lower extremity with ulcer and inflammation | <0.4 | 0.0 | 0.2 | 0.2 | <0.1 | 0.2 |
| | vasculitis | 4.2 | 14.4 | 4.0 | 5.7 | 17.5 | 3.3 |
| | vasculitis of medium sized vessel | <0.4 | <0.6 | 0.1 | 0.1 | 0.2 | 0.1 |
| | vasculitis of the skin | 0.0 | <0.6 | 0.1 | 0.2 | 0.1 | 0.2 |
| **Pernicious anaemia** | pernicious anaemia | 0.0 | 0.0 | 0.0 | 0.5 | 0.0 | 0.0 |
| **Coeliac disease** | coeliac disease | 0.9 | <0.6 | 0.5 | 1.6 | 5.1 | 0.7 |
| **Scleroderma** | crest syndrome | <0.4 | 0.0 | 0.1 | 0.1 | 0.0 | 0.0 |
| | limited systemic sclerosis | <0.4 | 0.0 | 0.1 | 0.1 | 0.0 | 0.0 |
| | localized scleroderma | <0.4 | <0.6 | 0.1 | 0.5 | 0.3 | 0.1 |
| | systemic sclerosis | 0.6 | <0.6 | 0.2 | 0.4 | 0.8 | 0.2 |
| | systemic sclerosis with limited cutaneous involvement | <0.4 | 0.0 | 0.1 | 0.1 | 0.0 | 0.0 |
| **Sarcoidosis** | cardiac sarcoidosis | <0.4 | 0.0 | 0.0 | 0.1 | 0.0 | 0.0 |
| | cutaneous sarcoidosis | <0.4 | 0.0 | 0.0 | 0.1 | <0.1 | 0.2 |
| | lymph node sarcoidosis | 0.0 | 0.0 | 0.0 | 0.0 | <0.1 | 0.1 |
| | pulmonary sarcoidosis | 1.0 | 0.0 | 0.5 | 0.7 | 0.3 | 1.8 |
| | sarcoid heart muscle disease | <0.4 | 0.0 | 0.0 | 0.1 | 0.0 | 0.0 |
| | sarcoidosis | 2.7 | 0.0 | 1.0 | 2.1 | 1.1 | 2.6 |
| | sarcoidosis of lung with sarcoidosis of lymph nodes | <0.4 | 0.0 | 0.1 | 0.1 | <0.1 | 0.3 |
| **Ulcerative colitis** | abscess of intestine co-occurrent and due to ulcerative colitis | 0.0 | 0.0 | 0.0 | 0.0 | 0.0 | 0.1 |
| | chronic ulcerative colitis | 0.6 | 0.0 | 0.4 | 1.2 | 0.1 | 0.2 |
| | chronic ulcerative enterocolitis | 0.0 | 0.0 | 0.0 | 0.1 | 0.0 | 0.0 |
| | chronic ulcerative pancolitis | 0.6 | 0.0 | 0.3 | 0.9 | <0.1 | 0.2 |
| | chronic ulcerative rectosigmoiditis | 0.0 | 0.0 | 0.1 | 0.3 | <0.1 | 0.1 |
| | complication due to chronic ulcerative pancolitis | <0.4 | 0.0 | 0.1 | 0.2 | 0.0 | 0.1 |

| | | | | | | | |
|---|---|---|---|---|---|---|---|
| | complication due to chronic ulcerative rectosigmoiditis | 0.0 | 0.0 | 0.0 | 0.1 | 0.0 | 0.0 |
| | complication due to ulcerative colitis | 0.8 | 0.0 | 0.3 | 0.9 | <0.1 | 0.8 |
| | fistula of intestine due to ulcerative colitis | 0.0 | 0.0 | 0.0 | 0.0 | 0.0 | 0.1 |
| | intestinal obstruction due to ulcerative colitis | 0.0 | 0.0 | 0.0 | 0.0 | 0.0 | 0.1 |
| | left sided ulcerative colitis | <0.4 | 0.0 | 0.1 | 0.2 | <0.1 | 0.4 |
| | rectal hemorrhage due to chronic ulcerative pancolitis | <0.4 | 0.0 | 0.1 | 0.2 | <0.1 | 0.0 |
| | rectal hemorrhage due to chronic ulcerative rectosigmoiditis | <0.4 | 0.0 | 0.0 | 0.1 | 0.0 | 0.0 |
| | rectal hemorrhage due to ulcerative colitis | <0.4 | 0.0 | 0.2 | 0.5 | <0.1 | 0.7 |
| | ulcerative colitis | 1.9 | <0.6 | 1.3 | 2.5 | 4.1 | 2.6 |
| | ulcerative enterocolitis | 0.0 | 0.0 | 0.0 | 0.1 | 0.0 | 0.0 |
| | ulcerative pancolitis | 0.6 | 0.0 | 0.3 | 0.9 | <0.1 | 0.2 |
| | ulcerative proctocolitis | 0.0 | 0.0 | 0.1 | 0.3 | <0.1 | 0.1 |
| **Crohn's disease** | abscess of intestine co-occurrent and due to crohn's disease | <0.4 | 0.0 | 0.0 | 0.1 | 0.0 | 0.1 |
| | abscess of intestine co-occurrent and due to crohn's disease of large intestine | <0.4 | 0.0 | 0.0 | 0.1 | 0.0 | 0.1 |
| | abscess of intestine co-occurrent and due to crohn's disease of small and large intestine | <0.4 | 0.0 | 0.0 | 0.0 | 0.0 | 0.1 |
| | abscess of intestine co-occurrent and due to crohn's disease of small intestine | <0.4 | 0.0 | 0.0 | 0.1 | 0.0 | 0.1 |
| | complication due to crohn's disease | 1.7 | 0.0 | 0.6 | 1.8 | 0.1 | 1.1 |
| | complication due to crohn's disease of large intestine | 1.2 | 0.0 | 0.3 | 1.0 | 0.0 | 0.6 |
| | complication due to crohn's disease of small and large intestines | 0.9 | 0.0 | 0.1 | 0.5 | 0.0 | 0.3 |
| | complication due to crohn's disease of small intestine | 1.3 | 0.0 | 0.3 | 0.9 | <0.1 | 0.5 |
| | crohn's disease | 2.3 | <0.6 | 1.2 | 3.0 | 2.9 | 1.6 |
| | crohn's disease of intestine | 1.4 | <0.6 | 0.7 | 1.9 | 0.3 | 0.9 |
| | crohn's disease of large bowel | 1.0 | 0.0 | 0.4 | 1.3 | <0.1 | 0.6 |

| | | | | | | | |
|---|---|---|---|---|---|---|---|
| | crohn's disease of small and large intestines | 0.5 | 0.0 | 0.2 | 0.5 | 0.0 | 0.4 |
| | crohn's disease of small intestine | 1.0 | <0.6 | 0.5 | 1.2 | 0.2 | 0.7 |
| | fistula of intestine due to crohn's disease of small and large intestine | <0.4 | 0.0 | 0.0 | 0.1 | 0.0 | 0.1 |
| | fistula of large intestine due to crohn's disease | 0.0 | 0.0 | 0.0 | 0.2 | 0.0 | 0.1 |
| | fistula of small intestine due to crohn's disease | <0.4 | 0.0 | 0.0 | 0.1 | <0.1 | 0.0 |
| | gastrointestinal crohn's disease | 1.4 | <0.6 | 0.7 | 1.9 | 0.3 | 0.9 |
| | intestinal obstruction due to crohn's disease | <0.4 | 0.0 | 0.1 | 0.3 | <0.1 | 0.2 |
| | intestinal obstruction due to crohn's disease of large intestine | <0.4 | 0.0 | 0.1 | 0.2 | 0.0 | 0.1 |
| | intestinal obstruction due to crohn's disease of small and large intestine | <0.4 | 0.0 | 0.0 | 0.2 | 0.0 | 0.1 |
| | intestinal obstruction due to crohn's disease of small intestine | <0.4 | 0.0 | 0.1 | 0.3 | <0.1 | 0.2 |
| | rectal hemorrhage due to crohn's disease | <0.4 | 0.0 | 0.1 | 0.2 | 0.0 | 0.1 |
| | rectal hemorrhage due to crohn's disease of large intestine | <0.4 | 0.0 | 0.0 | 0.1 | 0.0 | 0.1 |
| | rectal hemorrhage due to crohn's disease of small intestine | 0.0 | 0.0 | 0.0 | 0.1 | 0.0 | 0.1 |

**Supplementary table 2. Prevalence of autoimmune diseases in the year prior to the index date in patients hospitalised with COVID-19**

The condition with the highest prevalence in most databases (highlighted in yellow) was reported as the prevalence for the respective autoimmune disease in Table 2 of the manuscript.

| **Autoimmune disease** | **Condition** | **CUIMC** | **HIRA** | **IQVIA-Open Claims** | **Optum EHR** | **SIDIAP-H** | **VA-OMOP** |
|---|---|---|---|---|---|---|---|
| **Type 1 Diabetes Mellitus** | acidosis due to type 1 diabetes mellitus | <0.9 | 0.0 | 0.1 | 0.6 | 0.0 | <0.2 |
| | disorder due to type 1 diabetes mellitus | 3.8 | 2.0 | 5.4 | 6.9 | 1.4 | 5.6 |
| | gangrene due to type 1 diabetes mellitus | 0.0 | 0.0 | 0.1 | <0.2 | 0.0 | <0.2 |
| | hyperglycemia due to type 1 diabetes mellitus | 2.7 | 0.0 | 2.4 | 3.7 | <0.6 | 2.0 |
| | hypoglycemia due to type 1 diabetes mellitus | <0.9 | 0.0 | 0.6 | 1.1 | 0.0 | 0.5 |
| | mild nonproliferative retinopathy due to type 1 diabetes mellitus | 0.0 | 0.0 | 0.2 | 0.2 | 0.0 | 0.3 |
| | moderate nonproliferative retinopathy due to type 1 diabetes mellitus | 0.0 | 0.0 | 0.1 | <0.2 | 0.0 | 0.2 |
| | neuropathic arthropathy due to type 1 diabetes mellitus | 0.0 | 0.0 | 0.1 | 0.2 | 0.0 | <0.2 |
| | neuropathy due to type 1 diabetes mellitus | <0.9 | 0.0 | 1.0 | 1.5 | <0.6 | 0.9 |
| | nonproliferative diabetic retinopathy due to type 1 diabetes mellitus | 0.0 | 0.0 | 0.3 | 0.4 | 0.0 | 0.6 |
| | peripheral circulatory disorder due to type 1 diabetes mellitus | <0.9 | <0.6 | 0.6 | 0.7 | 0.7 | 0.4 |
| | peripheral neuropathy due to type 1 diabetes mellitus | <0.9 | 0.0 | 0.8 | 1.2 | <0.6 | 0.8 |
| | polyneuropathy due to type 1 diabetes mellitus | <0.9 | 0.0 | 0.7 | 1.0 | <0.6 | 0.5 |
| | pre-existing type 1 diabetes mellitus | 0.0 | 0.0 | 0.1 | 0.3 | 0.0 | 0.0 |
| | pre-existing type 1 diabetes mellitus in pregnancy | 0.0 | 0.0 | 0.1 | 0.3 | 0.0 | 0.0 |
| | pregnancy and type 1 diabetes mellitus | 0.0 | 0.0 | 0.1 | 0.3 | 0.0 | 0.0 |
| | renal disorder due to type 1 diabetes mellitus | 1.4 | <0.6 | 1.5 | 2.4 | 0.0 | 1.1 |
| | type 1 diabetes mellitus | 4.8 | 1.5 | 7.5 | 7.5 | 4.4 | 5.3 |

| | | | | | | | |
|---|---|---|---|---|---|---|---|
| | type 1 diabetes mellitus uncontrolled | 0.0 | 0.0 | 0.0 | 0.8 | 0.0 | 0.0 |
| | type 1 diabetes mellitus with arthropathy | 0.0 | 0.0 | 0.1 | 0.2 | 0.0 | <0.2 |
| | type 1 diabetes mellitus without complication | 3.6 | 1.0 | 4.6 | 4.0 | 4.3 | 2.3 |
| | ulcer of lower limb due to type 1 diabetes mellitus | <0.9 | 0.0 | 0.4 | 0.7 | <0.6 | 0.7 |
| **Rheumatoid arthritis** | myopathy due to rheumatoid arthritis | 0.0 | 0.0 | 0.1 | 0.0 | 0.0 | 0.0 |
| | polyneuropathy in rheumatoid arthritis | 0.0 | 0.0 | 0.1 | <0.2 | 0.0 | 0.0 |
| | rheumatoid arthritis | 4.8 | 18.9 | 4.9 | 8.8 | 5.4 | 4.0 |
| | rheumatoid arthritis - ankle and/or foot | 0.0 | 0.0 | 0.1 | <0.2 | 0.0 | <0.2 |
| | rheumatoid arthritis - hand joint | <0.9 | 0.0 | 0.2 | 0.5 | <0.6 | <0.2 |
| | rheumatoid arthritis of knee | <0.9 | 0.0 | 0.1 | 0.2 | 0.0 | 0.2 |
| | rheumatoid arthritis of shoulder | 0.0 | 0.0 | 0.1 | 0.3 | 0.0 | <0.2 |
| | rheumatoid arthritis of wrist | 0.0 | 0.0 | 0.1 | <0.2 | 0.0 | <0.2 |
| | rheumatoid lung disease with rheumatoid arthritis | 0.0 | 0.0 | 0.1 | <0.2 | 0.0 | <0.2 |
| | seronegative rheumatoid arthritis | 1.3 | 2.3 | 0.9 | 1.3 | 1.4 | 0.5 |
| | seropositive rheumatoid arthritis | 1.8 | 5.0 | 1.9 | 3.6 | 4.5 | 1.9 |
| **Psoriasis** | arthritis mutilans | 0.0 | 0.7 | 0.1 | <0.2 | 0.0 | 0.6 |
| | guttate psoriasis | 0.0 | <0.6 | 0.0 | <0.2 | 1.0 | <0.2 |
| | psoriasis | 1.4 | 8.2 | 2.7 | 5.4 | 26.4 | 4.4 |
| | psoriasis vulgaris | <0.9 | 2.8 | 1.0 | 0.5 | <0.6 | 1.7 |
| | psoriasis with arthropathy | 0.0 | 0.7 | 0.6 | 1.7 | 2.5 | 0.9 |
| | psoriatic arthritis mutilans | 0.0 | 0.0 | 0.1 | <0.2 | 0.0 | 0.6 |
| | pustular psoriasis | 0.0 | 0.9 | 0.1 | <0.2 | 0.0 | <0.2 |
| **Psoriatic Arthritis** | arthritis mutilans | 0.0 | 0.7 | 0.1 | <0.2 | 0.0 | 0.6 |
| | psoriasis with arthropathy | 0.0 | 0.7 | 0.6 | 1.7 | 2.5 | 0.9 |
| | psoriatic arthritis mutilans | 0.0 | 0.0 | 0.1 | <0.2 | 0.0 | 0.6 |
| **Multiple sclerosis** | multiple sclerosis | 1.1 | <0.6 | 2.1 | 3.7 | 2.1 | 1.6 |
| **Systemic lupus erythematosus** | lung disease with systemic lupus erythematosus | 0.0 | 0.0 | 0.1 | <0.2 | 0.0 | <0.2 |
| | nephropathy co-occurrent and due to systemic lupus erythematosus | 1.1 | 0.0 | 0.3 | 1.0 | <0.6 | 0.2 |

| | | | | | | | |
|---|---|---|---|---|---|---|---|
| | nephrosis co-occurrent and due to systemic lupus erythematosus | 1.1 | 0.0 | 0.3 | 1.0 | <0.6 | 0.2 |
| | sle glomerulonephritis syndrome | 1.1 | 0.0 | 0.3 | 1.0 | <0.6 | 0.2 |
| | systemic lupus erythematosus | 3.2 | 1.7 | 1.9 | 4.3 | 2.6 | 0.9 |
| | systemic lupus erythematosus with organ/system involvement | 1.4 | 1.2 | 0.5 | 1.5 | <0.6 | 0.2 |
| **Hashimoto thyroiditis** | hashimoto thyroiditis | 0.0 | 0.0 | 0.0 | 0.9 | 0.0 | 0.0 |
| **Myasthenia gravis** | myasthenia gravis | <0.9 | <0.6 | 0.5 | 0.8 | 1.5 | 0.7 |
| | myasthenia gravis with exacerbation | <0.9 | 0.0 | 0.2 | 0.2 | 0.0 | 0.3 |
| | myasthenia gravis without exacerbation | <0.9 | 0.0 | 0.5 | 0.8 | 1.5 | 0.7 |
| **Vasculitis** | acute hemorrhagic gastritis | 0.0 | 6.9 | 0.4 | 0.4 | <0.6 | 0.5 |
| | antineutrophil cytoplasmic antibody positive vasculitis | <0.9 | <0.6 | 0.2 | 0.4 | <0.6 | <0.2 |
| | arteritis | <0.9 | <0.6 | 0.8 | 1.8 | 3.8 | 0.9 |
| | autoimmune vasculitis | 0.0 | <0.6 | 0.1 | <0.2 | <0.6 | <0.2 |
| | behcet's syndrome | 0.0 | 0.9 | 0.0 | <0.2 | <0.6 | <0.2 |
| | capillaritis | 0.0 | <0.6 | 0.1 | 0.0 | <0.6 | <0.2 |
| | deep thrombophlebitis | <0.9 | 1.6 | 0.3 | <0.2 | 8.6 | 0.4 |
| | deep vein phlebitis and thrombophlebitis of the leg | <0.9 | 1.6 | 0.3 | <0.2 | 8.6 | 0.4 |
| | granulomatosis with polyangiitis | <0.9 | 0.0 | 0.2 | 0.4 | <0.6 | <0.2 |
| | granulomatosis with polyangiitis with multisystem involvement | <0.9 | 0.0 | 0.1 | <0.2 | 0.0 | <0.2 |
| | hypersensitivity angiitis | 0.0 | <0.6 | 0.1 | 0.4 | <0.6 | <0.2 |
| | idiopathic capillaritis | 0.0 | <0.6 | 0.1 | 0.0 | <0.6 | <0.2 |
| | microscopic polyarteritis nodosa | 0.0 | <0.6 | 0.1 | <0.2 | 0.0 | <0.2 |
| | necrotizing vasculitis | 0.0 | <0.6 | 0.1 | <0.2 | 1.0 | 0.2 |
| | phlebitis | 1.8 | 4.7 | 2.8 | 4.7 | 15.5 | 2.3 |
| | phlebitis and thrombophlebitis | <0.9 | 1.6 | 1.0 | 1.3 | 11.9 | 1.1 |
| | phlebitis of deep veins of lower extremity | <0.9 | 1.6 | 0.5 | 0.3 | 8.6 | 0.5 |
| | phlebitis of lower limb vein | 1.1 | 3.0 | 2.2 | 3.5 | 12.2 | 1.6 |

| | | | | | | | |
|---|---|---|---|---|---|---|---|
| | phlebitis of superficial veins of lower extremity | <0.9 | <0.6 | 0.4 | 0.4 | 0.0 | <0.2 |
| | phlebitis of the femoral vein | 0.0 | 0.0 | 0.2 | <0.2 | 0.0 | <0.2 |
| | pigmented purpuric lichenoid dermatitis of gougerot and blum | 0.0 | <0.6 | 0.1 | 0.0 | <0.6 | <0.2 |
| | polyarteritis | 0.0 | <0.6 | 0.1 | 0.2 | 0.0 | <0.2 |
| | polyarteritis nodosa | 0.0 | <0.6 | 0.1 | 0.2 | 0.0 | <0.2 |
| | primary necrotizing systemic vasculitis | 0.0 | <0.6 | 0.1 | <0.2 | 0.0 | <0.2 |
| | primary systemic arteritis | 0.0 | <0.6 | 0.1 | 0.3 | 0.0 | <0.2 |
| | primary systemic vasculitis | <0.9 | 1.7 | 0.3 | 0.5 | 0.6 | 0.5 |
| | rheumatoid vasculitis | <0.9 | <0.6 | 0.1 | <0.2 | 0.0 | <0.2 |
| | secondary systemic vasculitis | <0.9 | <0.6 | 0.1 | 0.2 | <0.6 | <0.2 |
| | small vessel vasculitis | <0.9 | <0.6 | 0.3 | 0.4 | <0.6 | 0.2 |
| | small vessel vasculitis caused by immune complex | 0.0 | <0.6 | 0.1 | <0.2 | <0.6 | <0.2 |
| | systemic vasculitis | 1.1 | 2.6 | 0.4 | 0.7 | 0.6 | 0.5 |
| | temporal arteritis | <0.9 | <0.6 | 0.3 | 0.7 | 1.0 | 0.4 |
| | thromboangiitis | <0.9 | <0.6 | 0.0 | 0.0 | <0.6 | 0.2 |
| | thromboangiitis obliterans | <0.9 | <0.6 | 0.0 | 0.0 | <0.6 | 0.2 |
| | thrombophlebitis | <0.9 | 3.3 | 1.0 | 1.4 | 11.9 | 1.1 |
| | thrombophlebitis of deep veins of lower extremity | <0.9 | 1.6 | 0.3 | <0.2 | 8.6 | 0.4 |
| | thrombophlebitis of lower extremities | <0.9 | 2.2 | 0.3 | <0.2 | 8.6 | 0.4 |
| | varicose veins of lower extremity with inflammation | <0.9 | 1.2 | 1.4 | 2.8 | 3.7 | 1.0 |
| | varicose veins of lower extremity with ulcer and inflammation | <0.9 | 0.0 | 0.3 | 0.5 | 0.0 | 0.3 |
| | vasculitis | 3.4 | 14.4 | 4.4 | 7.7 | 20.8 | 4.4 |
| | vasculitis of medium sized vessel | 0.0 | <0.6 | 0.1 | 0.2 | 0.0 | <0.2 |
| | vasculitis of the skin | 0.0 | <0.6 | 0.1 | 0.2 | <0.6 | 0.3 |
| **Pernicious anaemia** | pernicious anaemia | 0.0 | 0.0 | 0.0 | 0.4 | 0.0 | 0.0 |
| **Coeliac disease** | coeliac disease | <0.9 | <0.6 | 0.3 | 0.9 | 1.2 | 0.4 |
| **Scleroderma** | crest syndrome | <0.9 | 0.0 | 0.1 | <0.2 | 0.0 | 0.0 |

| | | | | | | | |
|---|---|---|---|---|---|---|---|
| | limited systemic sclerosis | <0.9 | 0.0 | 0.1 | <0.2 | 0.0 | 0.0 |
| | localized scleroderma | 0.0 | <0.6 | 0.0 | 0.2 | 0.6 | <0.2 |
| | systemic sclerosis | <0.9 | <0.6 | 0.2 | 0.4 | 0.9 | <0.2 |
| | systemic sclerosis with limited cutaneous involvement | <0.9 | 0.0 | 0.1 | <0.2 | 0.0 | 0.0 |
| **Sarcoidosis** | cardiac sarcoidosis | <0.9 | 0.0 | 0.1 | <0.2 | 0.0 | <0.2 |
| | cutaneous sarcoidosis | 0.0 | 0.0 | 0.0 | <0.2 | <0.6 | 0.2 |
| | pulmonary sarcoidosis | 1.3 | 0.0 | 0.6 | 0.6 | 0.6 | 1.6 |
| | sarcoid heart muscle disease | <0.9 | 0.0 | 0.1 | <0.2 | 0.0 | <0.2 |
| | sarcoidosis | 3.4 | 0.0 | 1.2 | 1.9 | 1.2 | 2.1 |
| | sarcoidosis of lung with sarcoidosis of lymph nodes | <0.9 | 0.0 | 0.1 | <0.2 | <0.6 | 0.3 |
| **Ulcerative colitis** | chronic ulcerative colitis | 0.0 | 0.0 | 0.5 | 0.9 | <0.6 | <0.2 |
| | chronic ulcerative pancolitis | 0.0 | 0.0 | 0.4 | 0.7 | <0.6 | <0.2 |
| | chronic ulcerative rectosigmoiditis | 0.0 | 0.0 | 0.1 | 0.2 | <0.6 | 0.0 |
| | complication due to chronic ulcerative pancolitis | 0.0 | 0.0 | 0.1 | 0.2 | 0.0 | 0.0 |
| | complication due to ulcerative colitis | 0.0 | 0.0 | 0.3 | 0.7 | 0.0 | 0.6 |
| | left sided ulcerative colitis | 0.0 | 0.0 | 0.1 | 0.2 | <0.6 | 0.4 |
| | rectal hemorrhage due to ulcerative colitis | 0.0 | 0.0 | 0.1 | 0.2 | 0.0 | 0.4 |
| | ulcerative colitis | <0.9 | <0.6 | 1.3 | 2.2 | 2.8 | 1.6 |
| | ulcerative pancolitis | 0.0 | 0.0 | 0.4 | 0.7 | <0.6 | <0.2 |
| | ulcerative proctocolitis | 0.0 | 0.0 | 0.1 | 0.2 | <0.6 | 0.0 |
| **Crohn's disease** | abscess of intestine co-occurrent and due to crohn's disease | <0.9 | 0.0 | 0.1 | <0.2 | 0.0 | <0.2 |
| | complication due to crohn's disease | <0.9 | 0.0 | 0.5 | 1.5 | <0.6 | 1.0 |
| | complication due to crohn's disease of large intestine | <0.9 | 0.0 | 0.2 | 1.0 | 0.0 | 0.6 |
| | complication due to crohn's disease of small and large intestines | <0.9 | 0.0 | 0.1 | 0.4 | 0.0 | 0.3 |
| | complication due to crohn's disease of small intestine | <0.9 | 0.0 | 0.3 | 0.8 | 0.0 | 0.4 |

| | | | | | | | |
|---|---|---|---|---|---|---|---|
| | crohn's disease | 1.1 | <0.6 | 1.0 | 2.4 | 2.4 | 1.2 |
| | crohn's disease of intestine | <0.9 | <0.6 | 0.5 | 1.5 | <0.6 | 0.7 |
| | crohn's disease of large bowel | 0.0 | 0.0 | 0.3 | 1.2 | <0.6 | 0.4 |
| | crohn's disease of small and large intestines | 0.0 | 0.0 | 0.1 | 0.5 | 0.0 | 0.2 |
| | crohn's disease of small intestine | <0.9 | <0.6 | 0.4 | 1.0 | <0.6 | 0.5 |
| | fistula of large intestine due to crohn's disease | 0.0 | 0.0 | 0.0 | 0.3 | 0.0 | <0.2 |
| | gastrointestinal crohn's disease | <0.9 | <0.6 | 0.5 | 1.5 | <0.6 | 0.7 |
| | intestinal obstruction due to crohn's disease | <0.9 | 0.0 | 0.1 | 0.3 | <0.6 | 0.2 |
| | intestinal obstruction due to crohn's disease of large intestine | <0.9 | 0.0 | 0.1 | <0.2 | 0.0 | <0.2 |
| | intestinal obstruction due to crohn's disease of small intestine | <0.9 | 0.0 | 0.1 | 0.2 | 0.0 | <0.2 |
| | rectal hemorrhage due to crohn's disease | 0.0 | 0.0 | 0.1 | 0.4 | 0.0 | <0.2 |
| | rectal hemorrhage due to crohn's disease of large intestine | 0.0 | 0.0 | 0.0 | 0.2 | 0.0 | <0.2 |

**Supplementary table 3. Severe outcomes and mortality in 30 days post hospitalisation in patients with COVID-19 and prevalent autoimmune diseases**

| Outcome | Cohort | CUIMC | HIRA | IQVIA Open Claims | Optum EHR | SIDIAP-H | VA-OMOP |
|---|---|---|---|---|---|---|---|
| **Acute kidney injury** | COVID-19 | 9.9 | 2.8 | 16.7 | 22.2 | NA | 31.1 |
| | Influenza | 29.7 | 3.9 | 11.9 | 16.4 | 5.3 | 26.3 |
| **Acute myocardial infarction events** | COVID-19 | 2.5 | 2.6 | 2.4 | 6.3 | NA | 6.0 |
| | Influenza | 6.3 | NA | 3.4 | 4.1 | <1.5 | 7.8 |
| **Acute respiratory distress syndrome** | COVID-19 | 14.7 | 2.1 | 31.5 | 39.6 | NA | 42.8 |
| | Influenza | 17.1 | NA | 16.9 | 28.7 | 13.9 | 28.2 |
| **Cardiac arrhythmia during hospitalisation** | COVID-19 | 12.4 | 3.8 | 13.6 | 29.5 | NA | 35.1 |
| | Influenza | 24.6 | <3.3 | 17.0 | 28.1 | 19.2 | 32.0 |
| **Death** | COVID-19 | 24.6 | 6.3 | NA | NA | 18.0 | 16.3 |
| | Influenza | 3.4 | NA | NA | NA | 2.2 | 4.3 |
| **Heart failure during hospitalisation** | COVID-19 | 8.4 | 3.9 | 8.0 | 15.7 | NA | 24.5 |
| | Influenza | 35.4 | 5.2 | 14.1 | 20.5 | 14.9 | 28.3 |
| **Pneumonia during hospitalisation** | COVID-19 | 12.6 | 40.7 | 45.7 | 53.2 | NA | 33.0 |
| | Influenza | 28.0 | 25.5 | 30.1 | 36.3 | 22.6 | 19.5 |
| **Sepsis during hospitalisation** | COVID-19 | 4.7 | 4.9 | 17.3 | 23.5 | NA | 21.6 |
| | Influenza | 18.9 | <3.3 | 16.4 | 21.1 | 3.7 | 21.4 |
| **Stroke (ischaemic or haemorrhagic) events** | COVID-19 | 3.2 | 1.4 | 2.1 | 2.6 | NA | 3.4 |
| | Influenza | <2.9 | <3.3 | 2.0 | 2.6 | <1.5 | 2.4 |
| **Venous thromboembolic events** | COVID-19 | 3.2 | 1.4 | 3.3 | 7.6 | NA | 7.7 |
| | Influenza | 7.4 | <3.3 | 3.0 | 4.1 | 2.5 | 4.6 |
